# Global Epidemiology and Genetic Environment of *mcr* genes: A One Health Systematic Review of Current and Emerging Trends

**DOI:** 10.1101/2022.02.28.22271560

**Authors:** Masego Mmatli, Nontombi Marylucy Mbelle, John Osei Sekyere

## Abstract

**Background:** Mobile colistin resistance (*mcr*) genes modify Lipid A molecules of the lipopolysaccharide, changing the overall charge of the outer membrane.

**Methods:** A systematic review of all studies published between January 2015 to July 2021 was performed. Included articles described *mcr* genes in the context of their genetic environment, fitness cost, crystal structure, their enzymatic activity and the risk factors associated with the acquisition of *mcr*. Studies describing the epidemiology of *mcr* genes and novel therapeutics were included.

**Results and Discussion:** Ten *mcr* genes have been described to date within eleven Enterobacteriaceae species, with *Escherichia coli*, *Klebsiella pneumoniae,* and *Salmonella* species being the most predominant. They are present worldwide in 72 countries, with human specimens currently having the highest incidence. This is due to the wide dissemination of *mcr* in livestock animals, meat, manure, the environment, and wastewater samples, increasing the risk of transmission via foodborne, zoonotic, and vector-borne routes to humans. The stability and spread of *mcr* genes were mediated by mobile genetic elements such as the IncHI_2_ conjugative plasmid, which is associated with multiple *mcr*-variants and other antibiotic resistance genes. The cost of acquiring *mcr* is reduced by compensatory adaptation mechanisms. MCR proteins are well conserved via structurally. Hence, MCR-1 inhibitors and therapeutics should be applicable to all MCR proteins.

**Conclusion:** *Mcr* genes have spread from animals into the clinical setting, threatening public health. Combination therapies are a promising option for managing and treating colistin-resistant *Enterobacteriaceae* isolates whilst reducing the toxic effects of colistin.

**Importance/Highlights:**

- *Mcr* genes are associated with mobile genetic elements that are facilitating its global dissemination.
- Using colistin as a growth promoter increases the risk of acquiring *mcr-*positive *Enterobacteriaceae* in food-producing animals.
- There is a higher incidence of *mcr* genes in humans than animals.
- MCR proteins are phosphoethanolamine (PEtN) transferases that mediate the transfer of PEtN from its primary phosphatidylethanolamine to lipid A.
- Multiple compounds can synergistically restore colistin’s activity, reducing its dosage and toxicity.
- CRISPR-Cas9, endolysins-engineered enzymes, and antimicrobial peptides are promising therapies for colistin resistance

*Tweet: “Within bacteria, mobile colistin resistance genes have spread widely, with a high incidence in human samples. It’s critical to comprehend the genetic tools facilitating this spread and their mechanisms of action. These are detailed in this paper, as are new therapeutic strategies for managing these resistance genes.”*

## Introduction

Colistin was first introduced into clinical practice in the 1950s ^1^. It was derived from *Bacillus polymyxa* and belongs to polymyxins ^2^, a family of cationic polypeptide antibiotics with broad-spectrum antimicrobial activity ^3^. Colistin has a bactericidal effect on Gram-negative bacteria and thus is used for treating Gram-negative bacterial infections ^1, 2^. Cationic polypeptides have a high electrostatic attraction to the anionic lipopolysaccharide (LPS) located on the outer membrane of Gram-negative bacteria. There, it displaces the magnesium and calcium divalent cations (Mg^2+^ and Ca^2+^), which stabilise the LPS molecules ^1, 2^. This results in the disruption of the cell’s permeability, leading to cell death. However, due to the adverse side effects such as nephrotoxicity and neurotoxicity seen during colistin therapy ^2^, in the early 1980s, it was removed from human use and administered to food-producing animals as a growth promoter and therapeutics ^1, 4^. With the increasing resistance caused by Gram-negative pathogens such as carbapenem-resistant Enterobacteriaceae that threatens global public health ^5^, colistin was recently reintroduced as a last-line treatment option ^3, 4, 6^.

Colistin resistance was largely associated with chromosomal-encoded mechanisms that involved two component systems (TCSs) such as *pmrAB* and *phoPQ*, and mutation(s) in the *mgrB* regulator in *Klebsiella pneumoniae* ^7–9^. These chromosomal mutations mediated colistin resistance by modifying LPS, changing LPS’s overall charge, and reducing the affinity of polymyxins to the outer membrane ^7–9^. The types of modifications seen include the addition of phosphoethanolamine (PEtN) and 4-amino-4-deoxy-L-arabinose (Ara4N) to the 1-phosphate or 4-phosphate groups of Lipid A, respectively ^10^. The PEtN modification is associated with the *pmrAB* TCSs and the Ara4N modification with the *phoPQ* TCS alongside the *mgrB* regulator ^10^.

Plasmid-mediated colistin resistance (*mcr)* gene was first identified in both animals and humans by Liu *et al.* (2016) ^11^ from an *Enterobacteriaceae*. *mcr-1* increased colistin resistance and encoded a PEtN transferase enzyme that added PEtN to Lipid A ^11^ at the 4’-phosphate group ^12, 13^. The MCR-1 PEtN transferase enzyme had similar structural properties to EptC and LptA PEtN transferases from *Campylobacter jejuni* and *Neisseria meningitidis*, respectively ^14, 15^. The lipid A modification is identifiable with a matrix-assisted laser desorption/ionization-time of flight (MALDI-TOF) mass spectrometry (MS) assay with an additional *m/z*= 1920.5 peak observed in *mcr-*producing isolates, representing the modified Lipid A molecule ^16^. This activity is seen across the different *mcr* variants: *mcr-2* to *mcr-10*.

After the discovery of *mcr-1,* other novel *mcr* variants i.e., *mcr-2* to *mcr-10*, which are widely distributed within Enterobacteriaceae, have been reported globally. The identification of the *mcr-1* gene and its variants, *mcr-2* to *mcr-10*, is largely mediated by PCR screening and whole genome sequencing (WGS) tools, which are also used in *mcr* surveillance programmes ^11, 17–21^. Each *mcr* variant is mostly located on conjugative plasmids, associated with mobile genetic elements (MGEs), and mediates colistin resistance through PEtN transferase activity. *mcr* are widely distributed within Enterobacteriaceae, including *Escherichia coli, K. pneumoniae, Salmonella* species, and *Enterobacter* species, ^3, 11, 22, 23^ and have also been reported within other Gram-negatives such as *Pseudomonas aeruginosa* and *Acinetobacter* species ^24–26^.

Initially, food-producing animals were the reservoir of *mcr* genes due to the high usage of colistin in livestock ^11, 27, 28^. Farmers’ consumption of livestock and/or contact with livestock or their faeces was found to be a risk factor for infection with an *mcr*-producing isolate ^28, 29^. Global screening of *mcr-1* genes in livestock found increased number of *mcr-*producing isolates, resulting in a ban of colistin in food-producing animals for both growth promotion and treatment of bacterial infections ^30, 31^. The World Health Organisation (WHO), thereafter, listed colistin as part of the critically important antimicrobials for human medicine. This was to help preserve the effectiveness of colistin for clinical use and to minimize the transmission of *mcr* genes from animals, livestock, and the environment to humans ^30, 31^.

### Purpose of review

This review provides a map of the dissemination of *mcr-1* and its variants. It further evaluates the genomic content of each *mcr* variant, identifying the possible progenitor and the mobile elements that each is associated with. We largely look at the *mcr-1* gene, its structure and function, which enables it to mediate colistin resistance, the fitness cost imposed by *mcr-1* expression, and the risk factors enabling the dissemination of *mcr* genes. The review further summarizes the possible treatment options for *mcr-*producing colistin-resistant isolates.

Herein, we highlight the epidemiology and evolution of *mcr* genes over the last 6 years by providing insight into the genomic content of each variant, identifying MGEs that aid in its dissemination, their global distribution, the crystal structure, and enzymatic activity of MCR proteins, and promising emerging therapeutics that could manage *mcr-*positive *Enterobacteriaceae* infections.

## Methods

A comprehensive literature search was performed using PubMed. Journal articles published in English within the last six years (January 2016 to July 2021) were retrieved and screened using the following keywords: “colistin resistan*” or “polymyxin resistan*” and “mcr*” and “*Enterobacteriaceae*”. The search was focused on journal articles that discussed the crystal structure of MCR enzymes and their enzymatic activity, the genetic environment and genetic support (MGEs) of *mcr* genes, as well as their molecular epidemiology, risk factors and management. However, studies that involved reviews, books and documents, case reports, case studies were excluded. The inclusion and exclusion protocol used in this review is seen in Figure 1. The data that was extracted from the included studies is found in Table S1.

**Figure 1.**
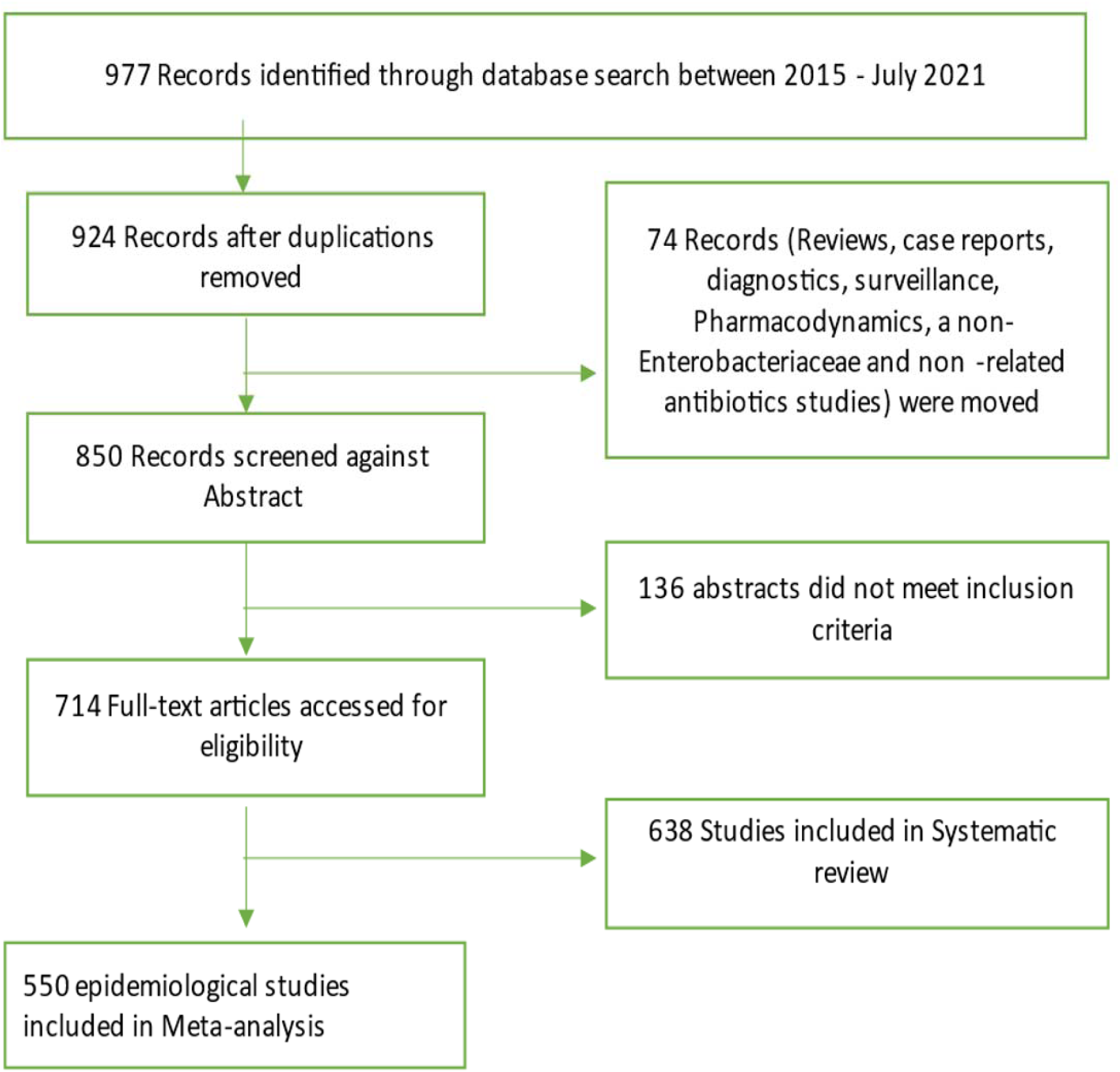
Flow diagram of literature search strategy and methodology. Out of 977 identified records between 2015 and 2021, a total of 638 studies were included in the systematic review whilst 550 of this were used for the epidemiological analyses.

## Results

### Characteristics of included studies

The study included 693 articles describing the epidemiology of *mcr* genes, the crystal structure and enzymatic activity of MCR proteins, the genomic environment of each *mcr* gene, the risk factors associated with the acquisition of *mcr*-positive *Enterobacteriaceae* isolates and the novel therapeutic options that can be explored for managing *mcr*-positive *Enterobacteriaceae*-related infections. A total of 429 articles were included in the epidemiological data and were used for the statistical analysis. The following data was extracted from the epidemiology articles: country, study year, specimen type and source, bacterial species, clone/MLST, *mcr*-type, mobile genetic elements (MGEs) and antibiotic resistance genes (ARGs) (Table S1).

### Enterobacteriaceae species distribution

*mcr* genes are well disseminated within Enterobacteriaceae, being frequently identified within *E. coli*, *K. pneumoniae* and *Salmonella* species, in that order. Other *Enterobacteriaceae* species that have been identified with *mcr* genes include *Cronobacter* species, *Citrobacter* species, *Kluyvera* species, *Leclercia* species, *Raoultella ornithinolytica,* and *Shigella* species (Figure 2A).

**Figure 2:**
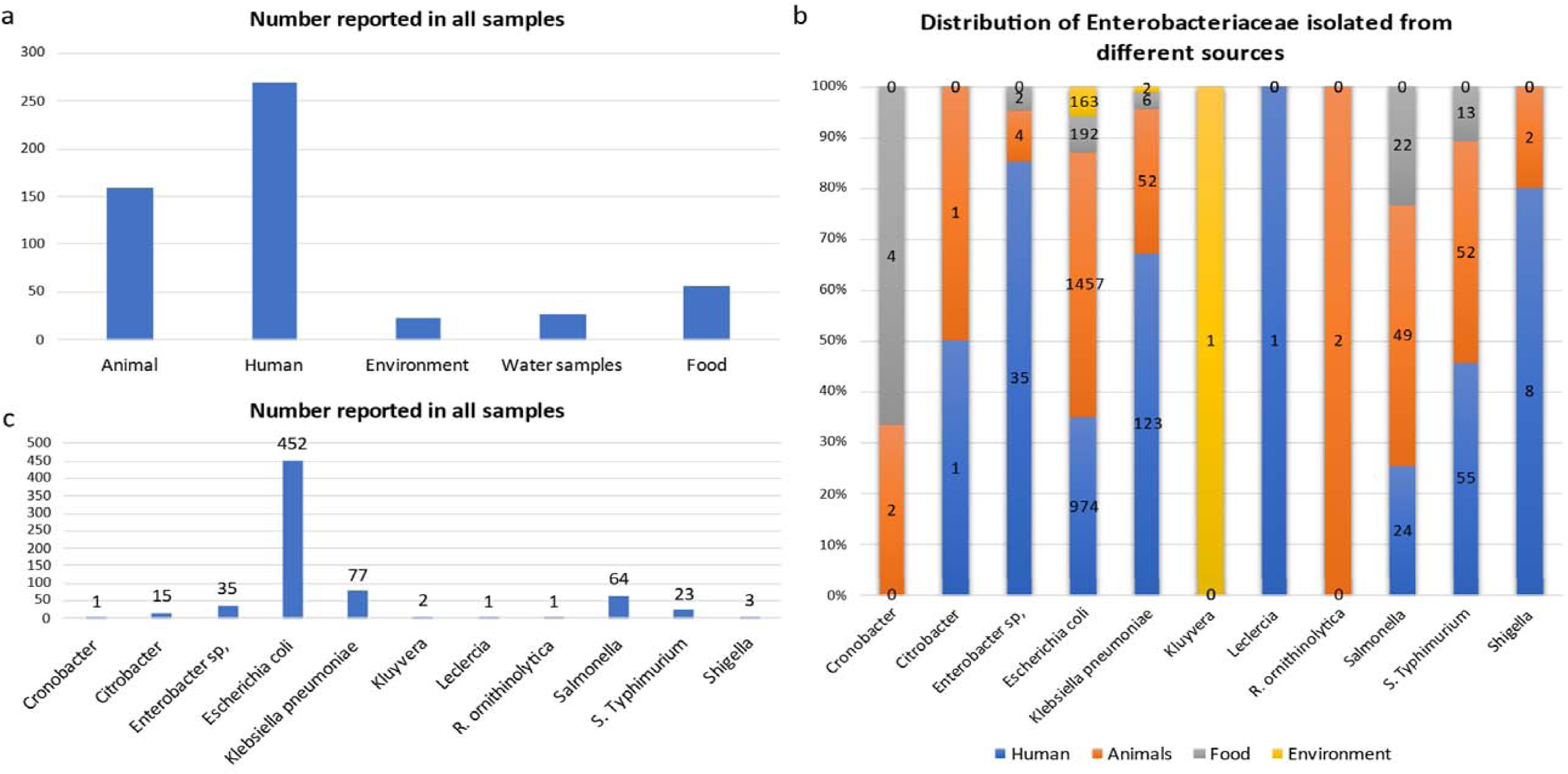
The sources and types of Enterobacteriaceae species identified to be harbouring *mcr* genes. A) Sources reported to harbour *mcr-*positive Enterobacteriaceae isolates were mainly animals, humans, water, food, and environmental samples. B) Distribution of *mcr-*producing Enterobacteriaceae species per sample source. C) The number of reports for each *mcr*-positive Enterobacteriaceae species.

Initially, *mcr* genes were commonly identified in animals. However, after evaluating the number of reports collected from 2016 to 2021, the most common source of *mcr*-positive *Enterobacteriaceae* isolates was human specimens (Figure 2a). In human specimens, *mcr* genes are frequently isolated from *Enterobacter* sp., *K. pneumoniae, Leclercia* sp., and *Shigella* sp (Figure 2b). *E. coli* is the most common *mcr*-positive isolate and is usually isolated from animal specimens (Figure 2c); this is also seen with *R. ornithinolytica*, *Salmonella* sp., and *S.* Typhimurium (Figure 2b). Other sources identified include food and the environment, which are mostly made up of wastewater samples. In the environment, *mcr-*positive isolates comprised only three species: *E. coli, K. pneumoniae,* and *Kluyvera* sp (Table S2). Lastly, *mcr*-positive species have been also isolated from food animal and vegetable samples, being isolated from mostly packaged meat and vegetables. These species include: *Cronobacter* sp., *Enterobacter* sp., *E. coli, K. pneumoniae, Salmonella,* and *S.* Typhimurium.

*Mcr-1* and its variants are the most common and well disseminated of all *mcr variants* (Figure 3a). This was later shown to be because of multiple factors such as fitness cost and MGEs. *Mcr*-1 has spread globally and has been identified in 69 countries, with *mcr-1* being the most prevalent *mcr* variant in most countries (Figure 3c). Although the other variants have spread globally, they have been identified in low numbers compared to *mcr-1* (Figure 3a). This includes *mcr-3,* which has been identified in fifteen countries, with Colombia and Thailand having the highest counts*. Mcr*-3 variants *(mcr-3.1*, *mcr-3.5,* etc) have been identified in an additional three countries, and in a total of eight countries. This brings the total count of countries with *mcr*-3 genes to eighteen (Table S3, Figure 3c). Other well disseminated *mcr* variants in low numbers includes *mcr-5* and *mcr-9,* identified in ten countries. *mcr-8* has been reported in seven countries, *mcr-4* in five countries, *mcr-7* and *mcr-10* in two countries, and *mcr-6* in Thailand only.

**Figure 3.**
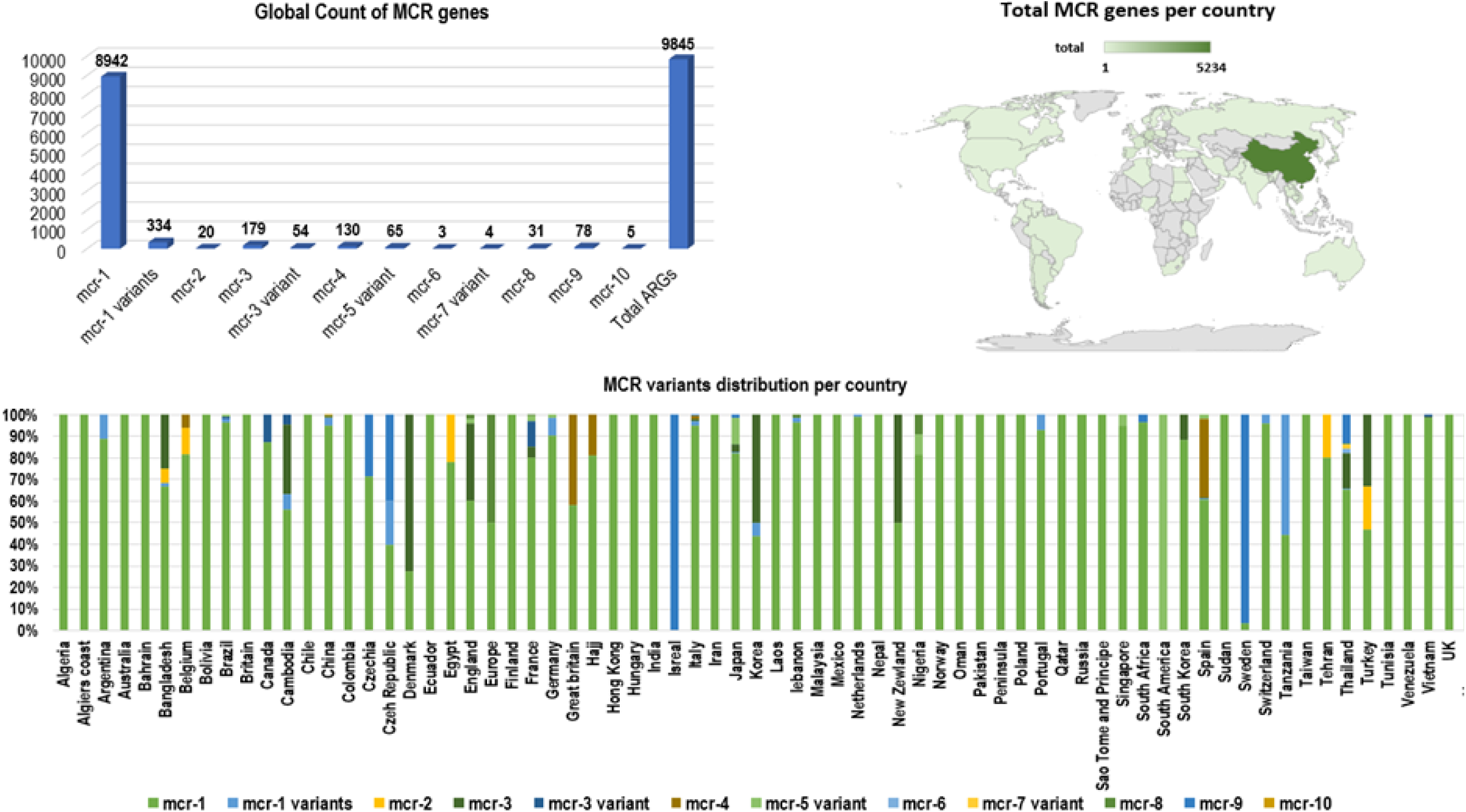
Global distribution and total count of mcr genes. A) Total number of mcr genes reported globally. B) The distribution of mcr genes per country. C) Global map showing the geographical distribution of mcr genes.

Amongst the 70 countries identified with *mcr* genes, countries such as Thailand harboured six *mcr-*variants (*mcr-1, mcr-3.mcr-6, mcr-7, mcr-8* and *mcr-9*) and China harboured eight variants (*mcr-1, mcr-3, mcr-4, mcr-5, mcr-7, mcr-8, mcr-9* and *mcr-10).* Other countries such as the USA, Turkey, Spain, Nigeria, Korea, Japan, Italy, France, England, Czech Republic, Cambodia, Brazil, Belgium, and Bangladesh harboured three to four *mcr-*variants, each inclusive of the *mcr-1* variants (Figure 3b, Table S3). Most *mcr* genes have been reported in studies from China (In Figure 3C), which is due to the large volumes of articles being published on *mcr* epidemiology from China.

It has thereafter been seen in China, that clones within *mcr-*positive *E. coli* (MPEC), which includes ST744, ST410, ST10, ST43, ST101 and ST206, have been identified in all four sources: food, environment, animals, and humans, seen in Table S4. These clones have, however, also disseminated globally, where ST744 has been identified in humans in eleven countries, in food in five countries, in animals in five countries and in the environment only in China. The MPEC ST10 strain is the most widely distributed within *E. coli*. It has been identified in animals in seventeen countries, humans in sixteen countries, the environment in five countries, and in food in five countries. MPEC clones within each country are usually found in both animals and humans, seen with ST744, ST69, ST117, ST131 and ST354 in Italy.

*S.* Typhimurium is also well distributed globally, with the *S.* Typhimurium ST34 strain identified in China, Colombia, Denmark, Germany, and the United Kingdom, in both animals and humans. In China, the *S.* Typhimurium ST34 strain has been identified in both humans and animals, and in food samples and human specimens in Germany. Similar results are seen in the monophasic variants of *S.* Typhimurium serovars (S.) 1,4,[5],12:i:-, and S. 4,[5],12:i:-, where ST34 was the only clone identified with *mcr* genes. The S. 1,4,[5],12:i:- ST34 strain has only been identified in Portugal in animals, food and humans, and the S. 4,[5],12:i:- has been identified in Belgium, Canada, Italy, Switzerland and the United States in animals and humans (Table S4).

The direct transmission of *mcr-*positive Enterobacteriaceae (MCRPE) isolates from animals and humans is discussed in this review (See Risk factors) and the data seen in Table S4 highlights this route of transmission.

### Geographical and host distribution of clones and ARGs

Similar to *mcr* genes (Figure 3c), other antimicrobial resistance genes (ARGs) are predominantly located in China (Figure 4), followed by Germany, Denmark, and England. Notably, several important ARGs are co-hosted by MCRPE isolates (Figure 3A). This is expected as colistin is used as a last-line antibiotic for multi-drug resistant (MDR) isolates. A Table showing the distribution of *mcr*-positive isolates hosting other ARGs is shown in Table S5.

**Figure 4.**
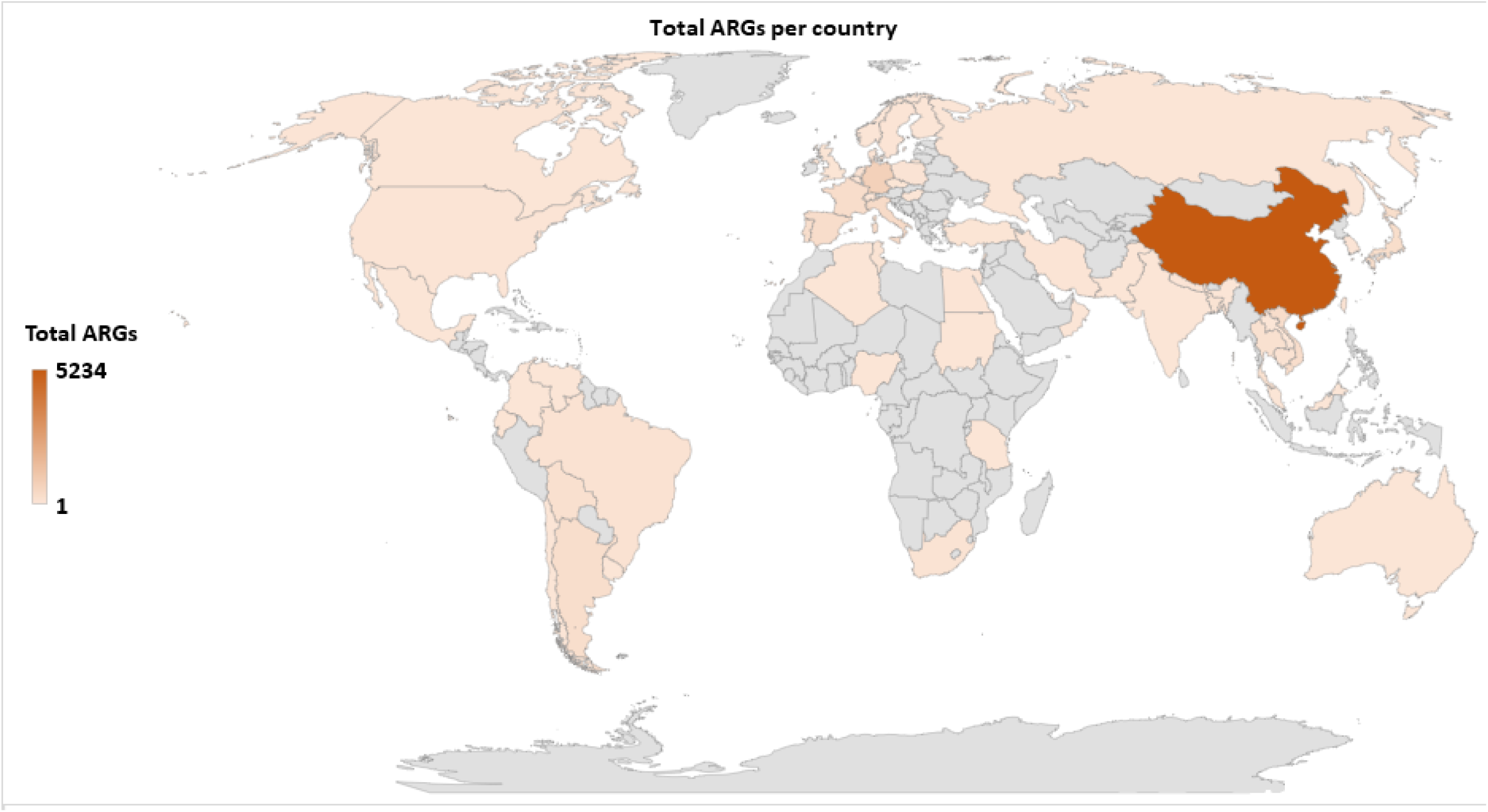
Global map showing the geographical distribution of antibiotic resistance genes found in all MCRPE.

### Plasmid incompatibility groups associated with mcr genes

*Mcr* genes are usually associated with insertion sequences (ISs), which aid in the mobilisation of resistance genes from the chromosome to plasmids and vice versa ^32^. The dissemination of *mcr* genes is, however, mediated by plasmids, which allow for the horizontal transfer and spread of resistance genes across different bacterial species, genera, and families ^33–35^. *Mcr* genes have been identified with 41 different plasmid replicons, where eight of these are made up of IncHI_2_ or IncHI_2A_. Most *mcr* variants are associated with multiple incompatibility groups, with *mcr-1* and its variants being associated with 37 plasmids; IncI_2_, IncX_4_ and IncHI_2_ were the most commonly reported (Figure 5). These incompatibility groups have also been shown to harbour other *mcr* variants: IncI_2_ has been associated with *mcr-7,* IncX_4_ with *mcr-2,* and IncHI_2_ with *mcr-*9, *mcr-3,* and its variants (Figure 5 and Table S6).

**Figure 5.**
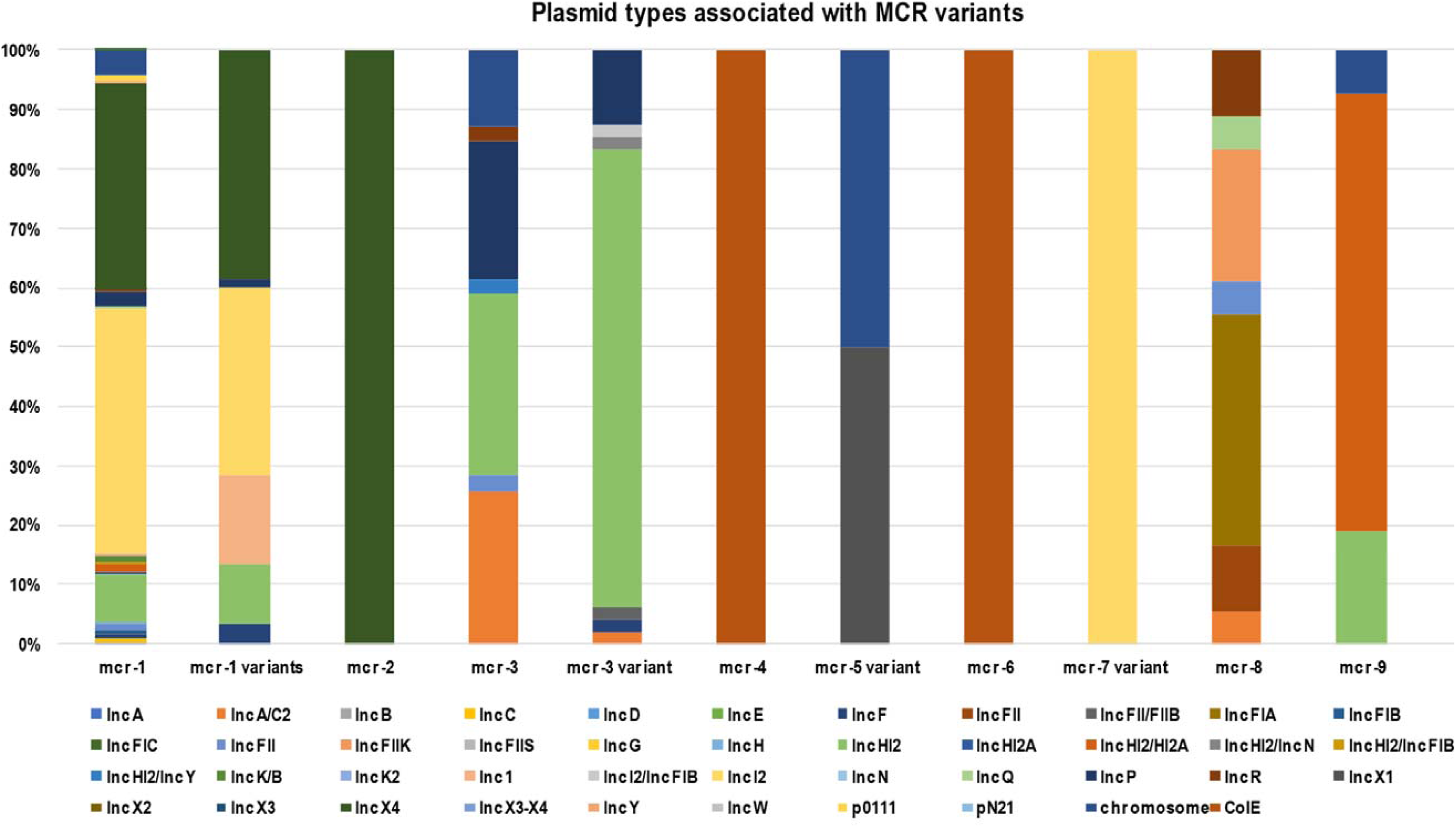
The distribution of plasmid incompatibility groups associated with each *mcr* gene.

Interestingly, the broad-spectrum IncHI_2_ plasmid was associated with the most *mcr* variants and is thus the main driver of *mcr* dissemination across different bacterial species. Further, *mcr-2*, *mcr-6,* and *mcr-7* have only been identified in a single incompatibility group (Figure 5). This is because these *mcr* variants have only been identified in low numbers. A heat map (Table S4) shows the MGEs profile of the included isolates, which includes both plasmid groups and IS elements.

## Discussion

Since the emergence of *mcr-1* in 2015, it has spread from animals to humans through several sources and routes, mediated by chromosomal and plasmid borne mobile genetic elements. The genetic context, associated risk factors, protein structure, and enzymatic activity of each *mcr* variant are discussed herein.

### Mcr-1 genomic content

The *mcr-1* gene is part of a 2,600 bp cassette that is made up of a putative promoter gene responsible for the expression of the *mcr-1* gene and the hypothetical protein later identified as pap2 ^36, 37^. *mcr-1* is speculated to have been derived from *Moraxella* species, which harbours the intrinsic chromosomal encoded *mcr*-like genes and the pap2 membrane-associated lipid phosphatase ^37, 38^. The *pap2* gene is found in both *mcr-1* and *mcr*-2 cassettes and shares 41% identity with *Moraxella oloensis* phosphatidic acid phosphatase ^20, 39^. *Moraxella mcr*-like genes with a significant degree of similarity to *mcr-1* and *mcr*-2 in *M. porci* and *M. osloensis* were respectively identified in Genbank. Thus, these genes could be closely related ^38^. Poirel *et al.* (2017a) identified an *mcr*-like gene, *mcr*-2.2, from an *M. pluranimalium* strain with an 82% and 99% amino acid identity to *mcr-1* and *mcr*-2, respectively.

An analysis of *mcr-1* sequences deposited in GenBank in 2017 revealed an *mcr-1*.10 variant from *Moraxella* sp. MSGI3-CO3 with 97.61% identity to the plasmid-borne *mcr-1* gene. This isolate was isolated from the faecal contents of healthy pigs in the United Kingdom in April, 2014 ^40^. This data suggests that *Moraxella* species may have been the likely source of *mcr-1* and *mcr*-2^37, 39^. Other evidence that supports the speculation that *mcr-1* evolved from the *Moraxella* species is the identification of an IS*Apl1* element in *M. bovoculi* and *M. porci* ^39, 41^. Li *et al.* (2018) suggest IS*Apl1* integrated into *M. bovoculi* and thereafter evolved with the *mcr*-like genes to the point seen today. This synteny of *mcr*-pap2 genes across *Moraxella* species further highlights this genus as a natural reservoir of *mcr*-like genes and a possible progenitor due to the high amino acid identity ^38, 39, 42^.

The mobilization of the *mcr*-pap2 unit was thereafter accomplished through IS elements, but Kieffer *et al.* (2017) identified a replicase gene associated with *mcr-1* on IncX_4_ plasmids. The gene had a 99% identity to *M. lacunata,* thus showing this species as a possible reservoir of IncX_4_ plasmids and further speculating that the *Moraxella* family may encode genetic tools likely involved in the initial mobilisation of *mcr* genes ^38^.

Stoesser *et al.* (2016) and Sun *et al.* (2018) suggested that the initial mobilisation of *mcr-1* genes into Enterobacteriaceae was IS*1294*-mediated, using a one-ended rolling circle transposition mechanism shown to be capable of mobilising adjacent sequences. The IS*1294* may have mobilised the *mcr-1* cassette into an IS*Apl1* composite transposon, creating an IS*Apl1*-*mcr-1*-pap2-IS*1294*-pap2-IS*Apl1* cassette, which has been identified on *E. coli* chromosome ^32, 43^. It was found that the cassette was still flexible enough to jump from the chromosome to a plasmid ^32^ by generating a putative circular intermediate product.

There is increasing evidence, however, that the *mcr-1* gene is mobilized primarily as a composite transposon, Tn*6330*, that is made up of two copies of IS*Apl1* that bracket cassettes ^44, 45^. IS*Apl1* is an IS that was first described in *Acinetobacillus pleuropneumoniae* and is part of the IS*30* family ^46^. The IS elements of this family are flanked by 20-30-base pairs (bp) inverted repeats (left IR (IRL), right IR (IRR)), which are essential for transposition ^47^. The IR contains a 924 bp open-reading frame that encodes a 44.3 kDa transposase protein containing a DDE domain, which encodes three conserved amino acid residues viz., D_228,_ D_295,_ and E_648_ (DDE), as well as carboxylase residues that help coordinate metal ions for catalysis ^44, 47^. Analysis of each IS*Apl1* element flanking the *mcr-1*-pap2 unit found conserved dinucleotides between the IS*Apl1* inverted repeats and the *mcr-1*-pap2 unit, an AT dinucleotide on the IRR of the upstream element and CG on the IRL of the downstream element ^44^. An interesting observation in the *mcr-1*.10 variant identified in *Moraxella* sp. MSGI3-CO3 was the presence of these dinucleotides, AT upstream and CG downstream, flanking the *mcr-1* structure ^40^. These dinucleotides were suggested to represent the ancestral target-site duplications (TSDs) formed during the initial mobilisation of *mcr-1* during IS*Apl1* insertion ^40^. Therefore, Snesrud *et al.* (2018) suggested that the formation of the composite transposon, Tn*6330*, was through two independent insertion events of IS*Apl1* into the TA-rich region of the *mcr-1*-pap2 unit, generating the conserved interior 2bp TSDs, AT and CG ^44^. Subsequent transposition of the Tn*6330* would therefore generate new TSDs at the new target site but would retain both internal conserved 2bp dinucleotides ^40^.

As stated above, the IS*Apl1* is part of the IS*30* family. The family has been shown to mobilize through a copy (out) and paste mechanism, forming circular intermediates of a single IS during transposition. The family is further known to have a high affinity for certain target sites resembling their IR sequence ^46–48^. Multiple studies have investigated the mechanisms of IS*Apl1* in mobilising the *mcr-1* gene and found that during each transposition event, the transposon was a circular intermediate covalently closed doubled stranded DNA, 5 699 bp in size and generated a 2 bp direct repeats at the insertion site which was an AT-rich region ^37, 44, 47, 49^, a similar mechanism seen in the IS*30* family. IS*Apl1* is most likely an important factor responsible for the insertion and fixation of the *mcr-1* gene into various classes of self-transmissible plasmids and host chromosomes ^37, 41, 50^. The formation of an intermediate structure consisting of IS*Apl1* and *mcr-1* during mobilization indicates that the resistance genes have become highly mobilizable in both plasmids and the chromosomes ^41^. After the first identification of *mcr-1* in pHNSHP45 ^11^, an Incl_2_ plasmid, *mcr-1* was thereafter detected in a wide range of conjugative plasmids, IncI_2_, IncHI_2_, IncX_4_, IncF, and IncP with the potential to mediate the dissemination of *mcr-1* genes into other Gram-negative bacteria ^50^. Petrillo *et al*. (2016) suggest that the insertion of the complete transposon triggers the rapid mobilization of conjugative plasmids, encouraging their dissemination across the *Enterobacteriaceae* family ^51^. The copy out and paste in mechanisms allow for the transposition of the *mcr-1* cassette, and the decay properties of IS*Apl1* further transfix the resistance gene into a plasmid or chromosome^49, 52^.

Snesrud *et al*. (2017) and Li *et al.* (2019) discovered that IS*Apl1* is highly active and that a single copy of IS*Apl1* can mobilize independently of *mcr-1* across the host genome in AT regions with a slight central GC bias ^49, 53^. A sequencing analysis of four *mcr-1* containing isolates performed by Snesrud *et al.* (2017) identified two to six copies of IS*Apl1* element throughout the isolates’ genome. The highly active nature of the IS*Apl1* elements thereafter triggers the deletion of the flanking IS*Apl1* copies to prevent further plasmid rearrangements ^40, 49^. Snesrud *et al.* (2016) analysis of the *mcr-1* sequence environment showed that Tn6330 has a strong tendency to decay through deletion, removing parts of, or both copies of IS*Apl1*, thus transfixing *mcr-1* into a vector plasmid. This has led to the observation of many sequences lacking one or both of IS*Apl1* ^37, 44^. Composite transposons in the IS30 family have been shown to contribute to replicon stabilization through transposition and illegitimate recombination ^44, 47^. The loss of IS*Apl1* elements results in the loss of transposability, stabilizing the *mcr-1* cassette in plasmids, which facilitates the widespread dissemination of the colistin resistance gene in self-transmissible plasmids ^44, 54, 55^. As discussed, IS*Apl1* has a significant bias for insertion in AT-regions and generates TSD of two or three bases. The analysis of the *mcr-1* cassette in the absence of IS*Apl1* elements has shown that the cassette is found in similar locations as per plasmid type and flanked by conserved trinucleotides (5’-ATA-3’) that are found immediately downstream of the IS*Apl1* IRR ^44, 56^. This is because the deletion event involves 1-4 flanking nucleotides that remain at the deletion junction ^40^.

An analysis of *mcr-1* sequences deposited in the public database has shown four general structures of *mcr-1* sequences: the complete composite transposon with both copies of IS*Apl1* elements; structures with a single copy of IS*Apl1* located downstream of *mcr-1*; structures that lost both copies and a rare fourth structure with a single copy of IS*Apl1* located upstream ^40, 44^. In structures with a single copy of IS*Apl1* found upstream and the IRR sequence of the downstream of deleted IS*Apl1*, Snesrud *et al.* (2016) suggest that the transposase encoded by the upstream IS*Apl1* can recognize the downstream IRR and thereafter still be able to mobilize the bracketed region without a complete composite transposon ^44^. The partially or complete removal of IS*Apl1* was through an illegitimate recombination that generated mismatches and deletions. Sun *et al.* (2018) found that the 3’ end of the *mcr* cassette unit was flexible in all IncX_4_ plasmids and the sequence could match with the perfect IRR of IS*Apl1*, though all IncX_4_ currently lack IS*Apl1* elements. The evidence from this study suggests that the TSD generated, and the six mismatches acquired through illegitimate recombination could be identified as a “relic” to track an insertion event that resulted in the subsequent loss of IS*Apl1* ^32^. The loss of IS*Apl1* elements in IncX_4_ was conducive to maintaining the *mcr-1* cassette on the plasmid ^32^, increasing its stability, and thus allowing for the dissemination of resistance genes via the plasmids ^44, 54, 55^. The identification of these IRR in IncX_4_ plasmids allows for the conclusion that IS*Apl1* was associated with the transposition of the *mcr-1* cassette into IncX_4_ plasmids ^57^.

### Fitness cost of mcr-1 genes on bacterial host

The acquisition of *mcr-1* bearing plasmids has been shown to have a beneficial effect on the host, improving bacterial survival in the presence of colistin treatment ^58^. The acquisition and expression of the *mcr-1* gene results in the incorporation of *MCR-1* into the bacterial membrane and the phosphoethanolamine (PEtN) modification of the lipopolysaccharides (LPS) ^59^. However, multiple studies have shown that the expression of *mcr-1* imposes a fitness cost by placing an energy burden on the host ^60^, impairing cell growth, and diminishing bacterial fitness ^59^. Andersson *et al.* (2006) ^61^ explained that a significant fitness cost is seen when the susceptible strain outcompetes the resistant strain in an antibiotic free environment. The imposed fitness cost of a conjugative plasmid harbouring a resistance gene has been shown to be because of various factors such as the resistance mechanisms, the bacterial species, and the antibiotic ^58^.

Evaluating the cost of colistin resistance on resistant strains, it is shown that certain chromosomal mutations within genes such as *mgrB* have no significant fitness cost on *K. pneumoniae* ^62^ whilst genes such as *pmrB,* do ^63^. Giordano *et al.* (2019) ^63^ found that the expression of the *mcr-1* gene imposes less of a burden on *K. pneumoniae* than the mutated *pmrB* gene. The chromosomal mutations within *pmrAB, crab, phoPQ,* and *mgrB* genes have been shown to mediate colistin resistance through the phosphorylation of lipid A ^64–66^. Tietgen *et al.* (2018) discovered that the fitness costs imposed by *mcr-1* plasmid carriage, such as growth rates and cytotoxicity, could be species-specific ^67^. Particularly, the acquisition of an *mcr-1*-harbouring plasmid in *K. pneumoniae* has been shown to impose a significant fitness cost on the host ^58^, but multiple studies have shown that the acquisition of an IncI_2_ plasmid, in various sizes, carrying *mcr-1* had no significant cost on the host ^68, 69^. This may be plasmid and/or species specific or may be due to acquired compensatory mutations within the IncI_2_ plasmid. However, the overexpression of *mcr-1* was seen to result in profound changes in the architecture of the outer membrane, resulting in the loss of membrane structural integrity and causing leakage of cellular cytoplasm, resulting in cell death ^59, 70^.

Yang *et al.* (2017a) ^59, 70^ found that the overexpression of *mcr-1* imposes a significant cost by decreasing the growth rate, causing significant membrane degradation and moderate fitness loss. They came to this conclusion through evaluating the effects of *mcr-1* expression on the relative fitness of *E. coli* TOP10 and found that with increasing levels of *mcr-1* expression resulted in a significant fitness burden on the host ^59, 70^. An analysis of the cellular morphology in the *mcr-1* overexpressed strains using a transmission electron microscopy showed cell architecture alterations and a complete loss of cellular morphology. The overexpression of *mcr-1* altered the structural integrity of the outer membrane and further impaired the cell membrane ^59^. They finally found that the embedding of *MCR-1* into the outer membrane and the PEtN modifications of Lipid A were the leading factors contributing to fitness cost and membrane degradation. Therefore, the expression of *mcr-1* in host strains is tightly controlled to regulate the *mcr-1* fitness cost ^70^.

*Mcr-1* expression is therefore quite toxic for the bacterial host, and the acquisition of a *mcr-1*- bearing plasmid thereafter results in compensatory adaptation that allows for the maintenance of high-cost conjugative plasmids ^19, 71, 72^. Dahlberg *et al.* (2003) ^72^ found that the cost of plasmid carriage is reduced over long-term culture because of compensatory mutations. The host chromosome or plasmid evolves compensatory mutations that, in the case of plasmids, enhance the fitness of the host, and in the case of chromosomal mutations, aid the host in evolving towards new growth conditions by decreasing the plasmid carriage cost ^72^. This has been seen in multiple studies where *mcr-1* bearing plasmids initially imposed a biological cost on the transformant. However, overtime, the cost of plasmid carriage in long-term cultures was largely compensated for and plasmids were stably maintained through passages ^59, 63, 67, 71, 73^.

Ma *et al*. (2018a) ^71^ investigated the potential mechanisms involved in compensatory adaptation through comparative genomics and identified single nucleotide polymorphisms in several genes. Amongst the genes identified in this study are *dnaK,* which encodes a molecular chaperon involved in chromosomal DNA replication, and *cpoB,* which encodes an RHS repeat protein involved in maintaining the cell envelope integrity during cellular division ^71^. These two genes are located on the chromosome and have non-synonymous single nucleotide polymorphisms (SNPs). The role of these genes in reducing plasmid carriage cost is unknown and may represent novel mechanisms. The study further shows that bacteria may use different strategies to reduce the fitness cost of plasmid carriage under different environmental conditions ^71^

In the absence of antibiotics (colistin) selective pressure, *mcr-1* bearing plasmids have been shown to be less maintained and that the complete elimination of the *mcr-1* gene within a population is possible ^58^. Nang *et al*. (2018) ^58^ performed a plasmid stability assay and found that in the absence of colistin, there was a gradual loss of the *mcr-1* plasmid, there was a decrease in its maintenance within the population. This may be due to the instability of *mcr-1* harbouring plasmids ^58^. Arcilla *et al*. (2016) have shown the complete elimination of *mcr-1* bearing bacteria in travellers returning to their home country after a month ^74^.

### MCR-1 structure

*MCR-1* is a 541 amino acid, integral membrane protein made up of two domains: a C-terminal periplasmic catalytic domain and an N-terminal 5’-helix transmembrane domain ^75, 76^. The transmembrane (TM) domain is made up of 5-membrane spanning α-helixes with an overall positive charge because of positively charged residues, which interact with the negatively charged phospholipid head groups of the membrane bilayer. The TM domain anchors the protein into the inner membrane and is connected to the catalytic domain through a bridging helix ^75^. The catalytic domain is made up of both positively and negatively charged residues, where the negatively charged residues create the binding pockets and allow them to be buried within the domain ^75^.

The overall shape of *MCR-1* is a hemispherical shape composed of several β-α-β-α motifs made up of β-strands sandwiched between α-helical structures ^12, 77^. The catalytic domain has an alkaline phosphatase family α/β/α fold, also with a hemispheric shape equipped with a zinc binding pocket containing a conserved phosphothreonine-285 residue (Figure 6)^12, 77^. The pocket is common to PEtN transferases and alkaline phosphatase; each enzyme, however, differs in the orientation and number of zinc molecules ^77^. The pocket is proposed to be critical for the nucleophilic attack of the phosphate of the donor PE substrate by stabilizing the alkoxide of the Thr285 ^77^. The Thr285 residue, is the catalytic nucleophile that acts as both a nucleophile and a PEtN acceptor during the catalytic mechanisms. The residue is critical to *MCR-1*’s function as mutations to this residue significantly decrease MCR-1’s activity ^77, 78^. The catalytic domain of MCR-1 is further made up of six cysteine residues that form three disulphide bonds, Cys281/Cys291, Cys356/Cys364 and Cys414/Cys422 ^77^ that stabilize and anchor the β-α-β-α motifs ^12^. These disulphide bonds are conserved, and equivalents are present in both LptA and EptC transferases ^77^, although LptA has four more cysteine residues.

**Figure 6.**
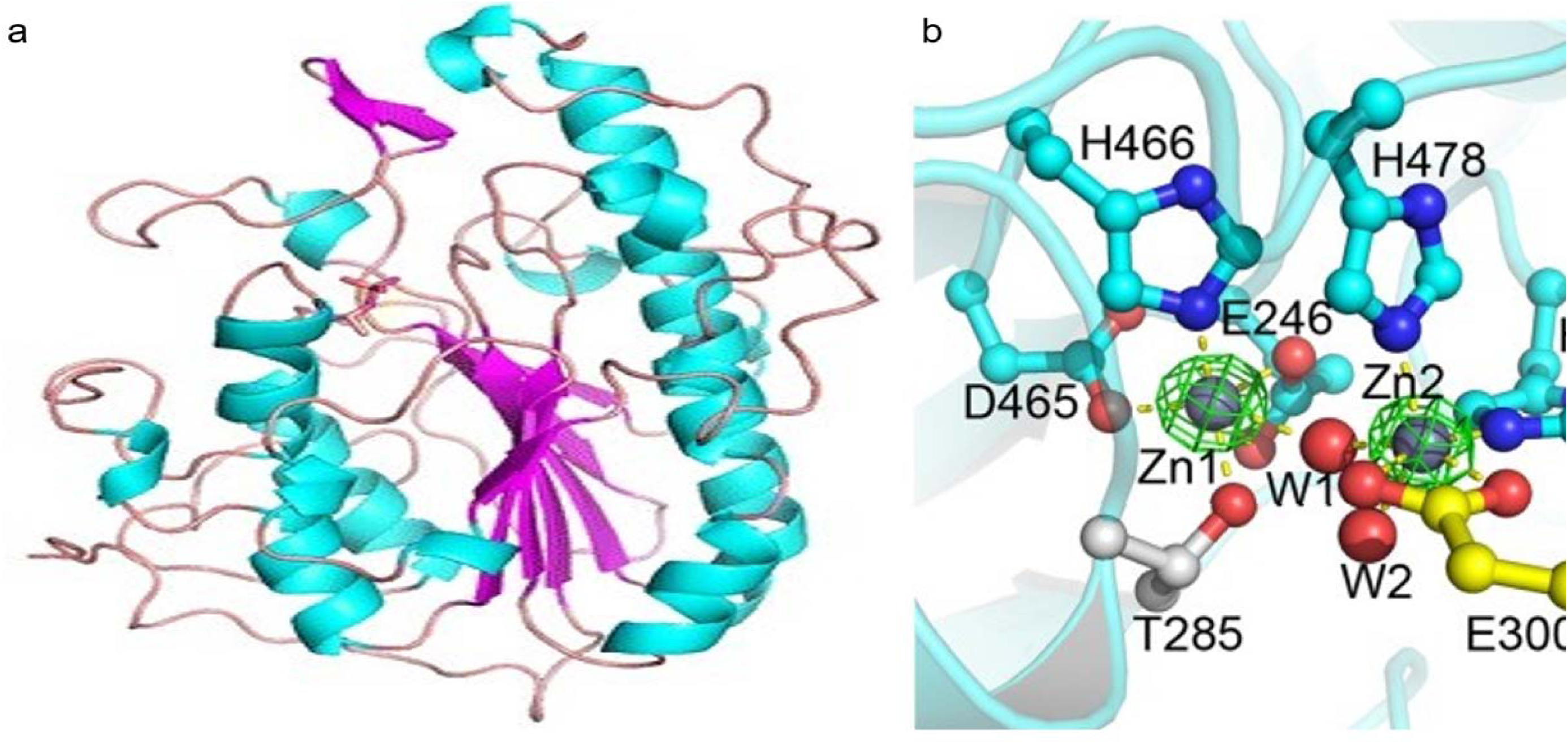
Structure of the catalytic domain of the MCR-1 protein and the five important residues required for its catalytic activity. A) The MCR-1’s hemispherical shape composed of the β-α-β fold made up of helices (cyan), strands (purple) and loops (salmon). Image was obtained from Stojanoski *et al.* (2016)^209^. B) A ball and stick model showing the conserved active site residues of MCR-1 consisting of Asp (D)465, Glu (E)246, His (H)466, His(H)395 and Thr (T)285, coordinated to the zinc ions (Zn1 and Zn2) and water molecules (W1 and W2). Image was obtained from Ma *et al.* (2016) ^210^.

The zinc binding pocket is in the active site of MCR-1 and is made up of conserved residues viz., Asp465, Glu246, His466, His395, and Thr285, and is made up of zinc molecules (Figure 6b)^77, 79^. The conserved residues are critical for the substrate binding of MCR-1 ^79^ and mutations in these amino acids abolish the MCR-1 activity, lowering the colistin MIC value down to control levels ^13^. The five conserved residues are well conserved in *MCR-1*, LptA, EptC and other alkaline phosphatase family members, although the nucleophile residue may differ between enzymes ^78^. MCR-1 has three zinc molecules clustered around the active site: Zn1 is buried within the active site and co-ordinated by the phosphate oxygen of Thr285; it is conserved in LptA and EptC ^77^. Zn2 is co-ordinated by His395, bound to a phosphate oxygen of Thr285 and three water molecules, forming a trigonal bipyramidal configuration seen in Figure 6b ^77^. Hu *et al.* (2016a) found that Zn2 is not critical for maintaining MCR-1 activity.

This may be because it is located on the surface of the active site. A structural comparison between LptA, EptC and MCR-1 found that the Zn2 site is less conserved. Hence, Hinchliffe *et al.* (2017) ^78^ concluded that an intact Zn2 site was not a prerequisite for MCR-1’s catalytic activity. However, His395 residue is important for the structure and activity of MCR-1. The Zn3 molecule, unlike the other zinc molecules, is not co-ordinated by the conserved residues but is tetrahedrally co-ordinated by four water molecules. The seven water molecules found in the active site, co-ordinating Zn2 and Zn3, are embedded in the protein through hydrogen bonding ^77^. The catalytic domain is thus a zinc-rich area (Fig. 6b), suggesting that MCR-1 may be able to attract zinc ions at different levels into the domain ^13^. The role of zinc in mediating lipid A modification, the catalytic mechanism, was evaluated by measuring the colistin MIC in the presence and absence of EDTA. Hinchliffe *et al.* (2017) ^78^ found clear reductions in colistin MIC values after EDTA treatment and concluded that the presence of zinc in the active site was important for MCR-1 function.

MCR-1 mediates the transfer of PEtN from the primary phosphatidylethanolamine (PE) to lipid A moiety at the 4’-phosphate group ^13, 79^. Wei *et al.* (2018) ^75^ and Liu *et al*. (2018) ^76^ investigated the molecular mechanism involved in this process. Wei *et al.* (2018) identified that the active site within the catalytic domain is made up of two binding pockets: the PEtN binding pocket and the Lipid A binding pocket. These binding pockets are in close proximity and are integrated within each other ^75^. Wei *et al.* (2018) ^75^observed that ethanolamine was a good analogue of PEtN and D-glucose for lipid A. Ethanolamine (ETA) is a good analogue as it mimics the covalent PEtN-enzyme intermediate. Lipid A is made up of two glucosamine units that are attached to acyl chains. Thus, monosaccharides and disaccharides can mimic a Lipid A molecule as they are both hexacyclic compounds ^75, 76^.

The PEtN binding pocket that accommodates ETA is made up of Glu246, Thr285, Asn329, Lys333, His395, Asp465, His466 and His478 residues ^75^. This pocket is responsible for the binding of phosphatidyl-ethanolamine (PE), which is stabilized through hydrogen bonding with Asn329, a water molecule, and phosphorylated Thr285 ^75^. Analysis of the entry of ETA into the pocket showed that the pocket undergoes conformational changes to adjust to the substrate. These included a 50° rotation of the His395, accomplished by breaking a hydrogen bond with a water molecule (wat7) and creating a new hydrogen bond with a new water molecule (wat8) ^75^. This releases the wat7 molecule from the PEtN binding pocket and thus, collectively, the conformational changes create room to accommodate the substrate, ETA. The entry of ETA further deprotonates the phosphorylated Thr285 residue, forming a PEtN molecule and a nucleophilic state at Thr285 ^75^. The Zn1 molecule subsequently facilitates the stabilization of the nucleophilic state of Thr285 that allows that residue to thereafter attack the PEtN, creating a PEtN enzyme intermediate. This constitutes the first step of the MCR-1-mediated catalytic process ^75^.

Wei *et al.* (2018) ^75^ and Liu *et al.* (2018) ^76^ investigated the mechanisms behind PEtN-transfer reaction with a lipid A analogue, D-glucose, identified within the Lipid A binding pocket. This pocket is made up of the following residues: Thr283, Ser284, Tyr287, Pro481 and Asn482, that is located near the Thr285 residue (Figure 7). The analysis of D-glucose found that D-glucose was held in the pocket by Thr285, Ser284 and Asn482 and was flanked by Tyr281 and Pro481 forming a sandwich structure of a π-π conjugative interaction ^75, 76^.The importance of the residues in both PEtN and Lipid A binding pockets in MCR-1 activity were evaluated through mutation construction. Mutations within Thr285, Asn329, Lys333, Glu246, His398, Asp465, His466 and His478 of the PEtN binding pocket and Tyr281 and Pro481 of the Lipid A binding pocket, resulted in a decrease in colistin MIC values ^75^. However, these mutations did not disrupt the protein expression and membrane localization of the MCR-1 protein. The mutations of Tyr281 and Pro481 in the lipid A binding pocket highlights the importance of this pocket in the MCR-1 activity. Wei *et al.* (2018) ^75^ suggests that it could bind to the glucosamine group of Lipid A and allows for the transfer of the reactive phosphate group from PEtN enzyme intermediate to lipid A. Thus, the second step of the catalytic process therefore involves the transfer of PEtN to Lipid A that is situated in a nearby pocket ^75^.

The MCR-1 catalytic domain is therefore made up of a PEtN binding pocket and a Lipid A binding pocket that mediate the transfer of PEtN from the PEtN binding pocket to lipid A moiety at the 4’-phosphate group ^13, 79^. This activity is validated through MALDI-TOF mass spectrometry (MS), provides *in-vivo* evidence of the MCR-1 activity ^79, 80^. This is accomplished by evaluating the LPS lipid A MS profile in the presence and absence of *mcr-1* expression as adding PEtN increases Lipid A mass units (+123) ^79, 81, 82^. In the absence of *mcr-1* expression, there is a single Lipid A peak (*m/z* = 1797.4) and in the presence of *mcr-1* expression, there are two unique peaks. The first peak is the wildtype bis-phosphorylated, hexa-acylated lipid A (*m/z* = 1797.4) and the second peak is the PPEtN-4’ (*m/z* = 1920.5) ^81^.

*MCR-1* has thus proven to have PEtN transferase activity and has a sequence and structural similarity to EptC and LptA PEtN transferases in *Campylobacter jejuni* ^14^ and *Neisseria meningitidis* ^83^, respectively ^77–79^. The active sites of the three PEtN transferases are highly conserved, i.e., the phosphorylated catalytic nucleophile Thr285, disulphide bonds, zinc binding pockets and the conserved active site residues ^78^. Therefore, these three sites are important for MCR-1 function; these conserved mechanistic activities and binding pockets will allow for the use of MCR-1 inhibitors to also inhibit chromosomally encoded PEtN transferases, restoring colistin susceptibility in acquired and intrinsically resistant bacteria ^77^.

### Mcr-2 variant

*Mcr*-2 was first identified in ten *E. coli* isolates during a passive surveillance screening of diarrhoea in calves and piglets, and like *mcr-1*, *mcr*-2 was located on a conjugative plasmid (IncX_4_) and was able to confer clinical colistin resistance (4-8 mg/L) ^20^. The gene was identified due to the presence of a putative membrane protein identified as a PEtN transferase in *mcr-1* negative isolates ^20^. This variant has a 76.75% and 80.65 % nucleotide sequence and amino acid identity, respectively, to *mcr-1* ^20^. *mcr*-2 encodes a 538 amino acid polypeptide consisting of two domains: a C-terminal periplasmic catalytic domain and an N-terminal 5’-helix transmembrane domain ^20, 33^. Sun *et al*. (2017b) found that the acquisition of the transmembrane domain allows for the correct localization of PEtN transferases within the periplasm and that the deletion of this domain impairs MCR-2’s ability to confer colistin resistance.

A genetic analysis of the IncX_4_ plasmid found that *mcr*-2 was located within two IS*1595*-like ISs and a 279 bp open reading frame (ORF), located downstream the *mcr*-2 gene ^20^. The ORF encodes a PAP2 membrane-associated lipid phosphatase which is related to the PAP2 protein encoded by the ORF of *mcr-1* ^20, 84^. The ORF of *mcr*-2 has a 41% identity to the phosphatidic acid phosphatase encoded by *M. osloensis*. When Xavier *et al.* (2016) removed the *mcr*-2-pap2 from the IS*1595* backbone, a BLASTn search produced a single hit to *M. bovoculi* strain with a 75% identity across a 100% query coverage. To support the hypothesis that the *mcr*-2 cassette originated from *Moraxella* sp., a phylogenetic analysis of the MCR-2 protein was performed and found that the protein was a distinct protein from *MCR-1* and might have evolved from *M. catarrhalis* ^20^. Sun *et al.* (2017b) suggests that the emergence of the PEtN transferases, *MCR-1*, MCR-2 and *Neisseria* LptA was a parallel evolutionary event for functional acquisition of colistin resistance during some environmental selection pressure, i.e., the intensive use of colistin in animal feed ^33^.

IS*1595* is composed of a transposase gene flanked by two inverted repeats (18 bp each). *Mcr*-2 is thus present within a complete transposon which allows for horizontal gene transfer ^33^. Sun *et al.* (2017b) found that the IS*1595* composite transposon was able to form circularized intermediates, which were identified during an inverse PCR assay. Hence, the IS*1595* elements are involved in the mobilisation of the *mcr-*2 cassette via homologous recombination events, which like *mcr-1,* allows for the dissemination of the resistance gene across diversified bacterial hosts ^33^.

After the first identification of *mcr*-2 in Belgium on an IncX_4_ plasmid, an effective vehicle for dissemination of resistance and virulence genes with a high transfer frequency ^20, 33^, it was thereafter identified in *Salmonella* sp. on IncX_4_ plasmids in Belgium in 2018 from retail meat collected in 2012 ^85^. It has subsequently been found in pig and poultry samples in 19 provinces in China, where the prevalence of *mcr*-2 in the study was 56.3% in poultry ^86^. *Mcr*-2 was then found in clinical settings in Iran, in stool samples collected between 2011-2016 ^87^. In Egypt, *mcr*-2 was widespread and identified in humans, wild birds, and the environment (water sources) ^88^. Compared to *mcr-1*, *mcr*-2 has not spread across the world and imposed a big threat to public health though found on a low fitness burden plasmid. The gene has however spread well within the *Enterobacteriaceae* family i.e., *E. coli, K. oxytoca*, *K. pneumoniae* and *Salmonella* sp.

### The mcr-2.2 variant: mcr-6

AbuOun *et al.* (2017)^42^ identified an *mcr-2.2* variant in *Moraxella pluranimalium* strain isolated from a pig in Great Britain. The MCR-2.2 protein was found to have an 87.9% amino acid identity to MCR-2, encoding 65 amino acid substitutions ^42^. Partridge *et al.* (2018) ^89^ suggested that since *mcr-2.2* encodes a protein that is 87.9% identical to the original MCR-2, it should be labelled as *mcr-6.* The *mcr-6* gene like *mcr-1* and *mcr*-2 was associated with the PAP2 gene located downstream of *mcr-6* ^42^. The *mcr-6* has only been identified in the *M. pluranimalium* MSG47-C17 strain isolated in 2012 and has not disseminated into the *Enterobacteriaceae* family. AbuOun *et al.* (2017) ^42^ however highlights that *Moraxella* species could have been the source of *mcr-pap2* genes.

### Mcr-3 variant

The first identification of *mcr-3* was in *E. coli* isolated from pigs in China ^90^. The identified 1626 bp putative PEtN transferase has a 45.0% and 47.0% nucleotide sequence identity to *mcr-1* and *mcr*-2 and an amino acid sequence identity of 32.5% and 31.7%, respectively ^90^. Using RaptorX, Yin *et al.* (2017) ^90^ predicted that MCR-3 protein, like MCR-1, and MCR-2, is made up of two Domains, domain 1 containing a five-transmembrane helix and domain 2 containing the periplasmic domain made up of the putative catalytic centre ^90^. Despite the difference between *mcr*-3 and the other *mcr* genes, Kieffer *et al.* (2018) ^91^ found that an *mcr*-3.12 variant had the same PEtN transferase activity and the expression of *mcr*-3 resulted in an 8-fold increase in MIC value of colistin ^92^. An LPS analysis showed that both MCR-1 and MCR-3 produced an identical additional peak at *m /z* 1921 in *mcr-1* and *mcr*-3 positive *E. coli*. Thus, the differences in nucleotide or amino acid sequences between the two *mcr* genes had no impact on the PEtN activity^91, 93^ and proving that MCR-3 confers colistin resistance in the same manner as MCR-1 and MCR-2 ^91^.

To further elaborate this observation, the expression of *mcr*-3 has been shown to significantly impair the cell wall integrity and decrease the electron density of *mcr*-3 producing *E. coli* ^94^, a characteristic that has been previously described in *mcr-1* producing isolates ^67, 70, 94^. Yang *et al.* (2020) ^94^ further evaluated the fitness cost imposed by *mcr-3* expression and compared it to that of *mcr-1* and found that the expression of both *mcr* genes had an impact on the bacterial fitness; however, the *mcr-3.1* and *mcr-3.5* imposed lesser fitness costs.

*Mcr*-3 genetic context is made up of a diacylglycerol kinase (*dgkA)* gene located downstream the *mcr*-3 gene and is thereafter flanked by a truncated (Δ) IS*Kpn40* upstream and an intact IS*Kpn40* downstream ^52^. In some cases, a ΔTnAs2 element is located upstream of the ΔIS*Kpn40* element ^52, 90, 95^, and the genetic context (ΔTnAs2-ΔIS*Kpn40*-*mcr-3*-*dgkA*IS*Kpn40*) is flanked by an IS26 element upstream and an intact IS*15DI* element downstream^96 34, 52^. These elements, IS*Kpn40*, TnAs2, IS*26* and IS*15DI* were identified and hypothesised to play a role in the transposition of the *mcr*-3 cassette among different species or bacterial genera ^35, 52, 97^. Sia *et al*. (2020)^52^ identified two circular intermediates of 3535 bp and 5990 bp made up of *mcr-3-dgkA-*IS*Kpn40* and ISΔ26-TnAs2-ΔIS*Kpn40*-*mcr-3-dgkA*IS*Kpn40*, respectively, during conjugation experiments. Wang *et al. (2018)* ^35^ evaluated the transferability of the 5990 bp circular intermediate and concluded that the ΔIS*26* and intact IS*15DI* mobilises the *mcr-3.1* via homologous recombination through the formation of circular intermediates. The IS-mediated transposition enables the mobilisation of the *mcr-3* resistance gene between the chromosome and plasmids, which further contributes to the dissemination of resistance genes ^34, 35^. Similar conclusion was reached with the 3535 bp circular intermediates in a study performed by Xiang *et al.* (2018), ^97^ which showed that mobilisation of the *mcr-3* core segment (*mcr-3-dgkA)* through IS*Kpn40* elements facilitated its spread into various plasmids.

The *mcr-3.12* variant has a 99% nucleotide sequence identity with an *Aeromonas veronii* sequence ^91^ and significant (77%) amino acid sequence identity to three PEtN transferases found within the genera ^90, 91, 98^. It is suggested that *mcr-3* genes originated from the *Aeromonas* genus, or the *mcr*-3 gene is widely disseminated as an acquired resistance trait and thus also found in *Aeromonads* ^91^. It is however thought that *mcr-3* was initially transposed from the *Aeromonas* genera to the *Enterobacteriaceae* family through IS*Kpn40*- mediated transposition. The IS*Kpn40* elements encode two ORF and the second ORF has a 99% identity to both a transposase from *Aeromonas sp.* and an integrase from *Aeromonas caviae* ^99^. This IS element is found flanking the *mcr-3* segment and was also shown to mobilise through homologous recombination ^52, 97^. The last evidence of *mcr-3* originating from *Aeromonas* includes the presence of Tn*As2* located upstream of the *mcr-3* gene ^90^. The transposon has only been identified in *A. salmonicida* and because its sequence in pWJ1 (original *mcr-3* plasmid) is a partial sequence, it is unlikely to have mobilised the *mcr-3* gene ^90, 95^. This evidence supports the possibility of the *Aeromonas* genus being a possible progenitor of *mcr-3* genes ^90^.

*Mcr-3* and its variants are disseminated within *Enterobacteriaceae*, having been identified in *E. coli, Salmonella sp.* and *K. pneumoniae* in healthcare centres, aquaculture, wastewater, and in animals ^18, 34, 90, 92, 95, 98, 100^. The resistance genes have been identified in both transmissible and non-transmissible plasmids such as IncA/C_2_, IncHI_2_, IncHI_2A_, IncF_II_/F_IB_, IncY, IncR, IncF, and IncP and in many variant mutations. The first identified *mcr-3* gene was identified on an IncHI_2_ plasmid replicon (pWJ1) in *E. coli* isolated from pigs in China ^90^. Thereafter, multiple *mcr-3* variants have been identified with one or more amino acid substitutions. Yang *et al.* (2020) evaluated the impact of *mcr-3* expression on bacterial fitness using two *mcr-3* variants, *mcr-3.1* and *mcr-3.5* (T488I, M23V, A456E) ^94^. The study showed that although the expression of both variants imposed a fitness cost and impaired the cell wall integrity, the expression of *mcr-3.5* was less costly ^94^. Yang *et al.* (2020) ^94^ showed that the amino acid substitutions A457V and T448I, seen in *mcr-3.5,* had strong compensatory effects when introduced in *mcr-3.1.* The introduction of these substitutions, individually, resulted in an increased fitness of up to 45%. However, double substitutions demonstrated a negative epistasis.

This study highlights an interesting concept about all *mcr* genes i.e., the *mcr* variants may encode compensatory mutations that mitigate the expression of *mcr* genes and thus, is an evolutionary mechanism for the worldwide dissemination of *mcr* genes ^94^. Though variants such as *mcr-3.10* has seven substitutions (V122G, R297L, E337K, H341Y, D358E, Q468K and I313V) ^92^ in both the putative transmembrane region and catalytic domains, these substitutions do not affect MCR-3.10’s ability to confer colistin resistance. Wang *et al.* (2018d) showed that similar to the original *mcr-3*, the expression of *mcr-3.10*, results in an 8-fold increase in colistin MICs. Similar results were seen with *mcr-3.5* ^95^.

### Mcr-4 variant

*Mcr-4* was first identified in *S.* Typhimurium isolated from the caecal content of a pig at slaughter in Italy in 2013 ^101^. The *S.* Typhimurium was colistin-resistant and negative for known *mcr* genes. MCR-4 respectively has an amino acid sequence identity of 34%, 35% and 49% to MCR-1, MCR-2, and MCR-3, and like other PEtN transferases, can mediate colistin resistance through Lipid A modification ^101^. MCR-4, however, has an 82%-99% amino acid sequence identity to one large PEtN transferase found in *Shewanella* sp., and the *mcr-4.3* variant has a 100% nucleotide sequence identity to a chromosomal PEtN transferase encoded by *S. frigidimarina* ^98, 101, 102^. *Mcr-3* and *mcr-4* are suggested to have originated from aquatic environments, as *Aeromonas* and *Shewanella* sp. are aquaculture fish pathogens that are intrinsically resistant to colistin ^98^. The dissemination of these *mcr* genes to the *Enterobacteriaceae* family may have been carried through plasmids from the host genome. The mechanism underlining this mobilisation is yet to be discovered ^98^.

*Mcr-4* was initially identified on an 8749 bp ColE10 plasmid, which encoded a RepB replicase, mobA/L mobilisation proteins, a RelE toxin and *excl1* gene ^101, 103^. In other cases, the ColE plasmid was found to encode a RepA replicase instead of RepB. However, both plasmids showed a 99% nucleotide sequence identity to plasmids from *Pantoea* species, with a 65% coverage ^101–103^. The *mcr-4* gene was flanked by IS5 elements on some plasmids and on other plasmids, it was flanked by an ΔIS10 element upstream and an IS1294 element downstream ^103^. Genetic analysis found that the ColE plasmid encodes a conserved 59-bp region upstream the *mcr-4* gene, and this region is predicted to encode the -35 (TTATTT) and -10 (AGCTAGTAT) promoter regions.^102^ This allows for ColE plasmids to replicate independently in different bacterial species and genera ^101^. ColE plasmids are broad-range non-conjugative plasmids whose mobilisation, however, require a helper plasmid; nevertheless, the mobilisation of the *mcr-4* gene is hypothesized to be achieved through a transposition event mediated by the IS5 element ^101^.

A genomic analysis of the initial *mcr-4-*producing *S.* Typhimurium genome found the ColE10 replicon located within the host chromosome integrated within the Type 1 methylation gene. Carattoll *et al.* (2017) suggest a transposition event mediated the chromosomal integration of the ColE10 plasmid ^101^. *Mcr-4* has been isolated from both humans and animals in Spain, Belgium, Italy, and China in *Enterobacteriaceae*^101, 104–106^. An inactive *mcr-4.3* variant identified by Teo *et al.* ^105^ (2018) was found to encode two missense mutations, V179G and V236F, that inactivated the PEtN transferase activity ^102, 105^. A comparative alignment of the MCR-4.3 to *MCR-1* and MCR-2 found that the active residues were conserved in MCR-4.3 ^105^. However, an MS spectrum of lipid A in *mcr-4.3-*expressing isolates showed *mcr-4.3* failed to modify lipid A ^102^. The transformation of *E. coli* with an *mcr-4.3* expressing vector resulted in no difference in colistin MIC values, suggesting that *mcr-4.3* does not confer colistin resistance ^102, 105^. Thus, some compensatory mutations in *mcr* may aid in fitness cost while others such as those seen in *mcr-4.3* significantly altered the PEtN transferase activity of *mcr-4.3* ^102^.

### Mcr-5 variant

*Mcr-5* was first identified in a d-tartrate-fermenting *S.* Paratyphi B isolated from food-producing animals across Germany between 2011 and 2013. Following this discovery of *mcr-5*, further screening resulted in the identification of fourteen additional *mcr-5*-producing isolates. These were isolated between 2011 to 2016 in the same study ^107^. *Mcr-5* encodes a 547 amino acid PEtN transferase, which has a protein identity of 33% to 35% to other known MCR proteins i.e., MCR-1 to MCR-4. Although MCR-5 has a low identity to other MCR proteins, all five MCR proteins, i.e., MCR1, MCR-2, MCR-3, MCR-4, and MCR-5, each encode the five conserved residue viz., E248, T286, H389, D458, and H359, that are located within the catalytic centre of the periplasmic domain ^107^. These conserved residues have been found to be essential for substrate binding and MCR activity ^79^. The capability of *mcr-5* to confer colistin resistance was shown using the original *mcr-5*-harbouring plasmid and a plasmid vector encoding the *mcr-5* operon in a transformation experiment using both an *E. coli* and a wild-type *S.* Paratyphi strains. In both experiments, the transformants achieved a colistin MIC value of 8 mg/L ^107^.

The MCR-5 protein has a 100% amino acid sequence identity to a PEtN transferase identified in *Pigmentiphaga* genus and *Cupriavidus gilardii.* The first 404 amino acids of the MCR-5 protein also have a high identity to a hypothetical protein from *Pseudomonas aeruginosa* ^107^. Genetic analysis of *mcr-5* found that the resistance gene is located within an operon encoding a Chromate B (ChrB) protein domain responsible for regulating the expression of ChrA for chromate resistance and two ORF that encode a major facilitator superfamily (MFS) type transporter ^107, 108^. The complete operon is located within a 7337bp Tn-*3* type transposon named Tn*6452* ^52, 107^, that is flanked by inverted repeats (IRs). A genetic analysis of the transposon identified *tnpA* encoding a Tn*3* transposon, *tnpR* encoding a Tn*3* resolvase and a *bla* gene encoding a β-lactamase ^109^. Tn*6452* encodes mechanisms for transposition of the *mcr-5* cassette, TnpA, and TnpR ^109^, which allows the *mcr-5* cassette to be transferred between plasmids and chromosomes ^110^.

*Mcr-5* has also been identified on Tn*3*-like structures that encode the ChrB gene but lacked the transposase gene and encoded an imperfect Tn*3*-like inverted repeats left (IRL) ^110–112^. This structure resembles features of a miniature inverted repeat transposable element, which possess a left or right IRs but lacks a transposase gene ^111^. Kieffer *et al.* (2019a) ^111^ suggests the acquisition of the *mcr-5* cassette was through a transposition event and although the Tn*3*- like structure lacks a transposase gene, it can still mobilise the cassette through a nonautonomous transposition mechanism. The *mcr-5* cassette has been identified on the host chromosome ^107^, a multicopy ColE-type plasmid ^107, 110, 112, 113^ and on an IncX plasmid ^110^. The presence of *mcr-5* cassette on a multicopy plasmid (ColE-type) thus means multiple copies of *mcr-5* transposon will exist within the cell. Borowiak *et al*. (2017) ^107^ found that there is a higher degree of colistin resistance (8 mg/L) in isolates harbouring ColE-type plasmids encoding *mcr-5* than in isolates with a single copy of the *mcr-5* integrated within the chromosome (4 mg/L). The ColE type plasmids are, however, non-conjugative and thus the presence of the *mcr-5* cassette on an IncX plasmid allows for horizontal transfer and dissemination of resistance gene between different bacterial genera and species ^109, 111^.

### Mcr-7 variant

*Mcr-7.1* was first identified in chicken in China ^114^. It has thereafter been identified in environmental samples ^115, 116^ and faecal samples in Brazil ^117^. The putative PEtN transferase has a 78% nucleotide sequence identity to part of the *mcr-3* gene ^114^. Genetic analysis of *mcr-7.1* identified a 381 bp ORF encoding the diacylglycerol kinase, *dgkA,* downstream of *mcr-7.1*. The *dgkA* has an 82% nucleotide sequence identity to *Aeromonas* ^114^. Yang *et al.* (2018) ^114^ suggests, like *mcr-3*, *mcr-7.1* originates from *Aeromanas*. This was derived based on the close genetic distance between MCR-3 and MCR-7.1 found during a phylogenetic analysis and the presence of *dgkA* gene downstream both *mcr-3* and *mcr-7.1.* The ORF sequence found upstream the *mcr-7.1* gene has an 81% nucleotide sequence identity to a putative phosphodiesterase gene found in *Aeromonas* ^114^. This evidence suggests that the *Aeromonas* genus is a possible progenitor of *mcr-7.1*.

The *mcr-7.1* encodes a 539 amino acid PEtN transferase that confers colistin resistance during conjugative experiments ^114^. The protein is made up of two domains, which resembles a similar structure of MCR-3. *Mcr-7.1* was identified on a self-transmissible IncI_2_ type plasmid with no ISs in its environment; hence, the dissemination of *mcr-7.1* may be achieved through plasmid mobilisation ^114^.

### Mcr-8 variant

*Mcr-8* was initially identified in *K. pneumoniae* from a swine faecal sample in China ^118^. Conjugation assays found that it is a functional PEtN transferase and its expression resulted in a four- and eight-fold increase in colistin MIC.^118–120^ MCR-8 mediated PEtN modification, which was identified using a MALDI-TOF-based technique that discriminates between colistin resistance mechanisms, i.e *mcr*-expression or chromosomal-mutations ^120^. BLASTn analysis showed that *mcr-8* has a 50.23% nucleotide and 39.96% amino acid sequence identity to part of *mcr-3* but has 30% to 40% amino acid sequence identity to other MCR proteins. Characterisation of MCR-8 using RaptorX predicted that MCR-8, like other MCR proteins, is made up of two domains: domain one consisting of a five transmembrane α-helix, and domain 2 consisting of the putative catalytic centre ^118^. Wang *et al.* (2018c) ^118^ performed sequence alignment of all eight MCR proteins and found that the active site residues (E246, T285, K333, H395, D465, H466, E468, H478) necessary for enzyme activity of MCR proteins and the six-cysteine residues required for the three disulphide bonds found in the catalytic domain were highly conserved across all 8 MCR proteins ^118, 119^.

Analysis of *mcr-8’s* genetic environment found that it is usually flanked by two complete IS*903* elements, with both 50 bp IRL and IRR located upstream, and downstream, respectively ^119, 121^. It is also associated with IS*Ecl1* and IS*Kpn26,* located upstream and downstream, respectively ^122^. Yang *et al.* (2019b) ^122^ found that the insertion of IS*Ecl7* was an independent event and may not be involved in mobilising *mcr-8*. The formation of circular intermediates via IS*903B* elements to mobilise *mcr-8* remains unknown; however, the association of *mcr-8* to ISs facilitates both the transmission of *mcr-8* and its close association with other resistance genes ^121^; specifically, it has been found associated with aminoglycoside resistance genes ^121^, β-lactamase genes ^119^ and *tmexCD1-toprJ1* genes that encode a novel plasmid-mediated efflux pump that confers resistance to tigecycline ^123^.

*Mcr-8* is commonly found in *K. pneumoniae* ^118, 120, 121, 123–126^, Wang *et al.* (2018c)^118^ performed a phylogenetic analysis of the *mcr-*positive *K. pneumoniae* isolates identified in their study and found that the *mcr-*positive *K. pneumoniae* were genetically diverse. *K. pneumoniae* express colistin resistance usually through chromosomal mutations ^10^. Wu *et al.* (2020) ^121^ suggest the expression of *mcr-8* may have synergistic effects with the chromosomal mutations within the two-component system (TCSs), resulting in heterogenous colistin resistance mechanisms ^121^. *Mcr-8* has also been identified in *Klebsiella quasipneumoniae* ^122^ and *R. ornithinolytica* as well as on a diverse range of Inc groups: IncFII, IncFIA, IncFIB, IncQ, IncR, IncFIIK, IncFIA and IncA/C; these will expectedly accelerate the dissemination of *mcr-8* and other ARGs within and outside *Enterobacteriaceae* ^119, 121^.

### Mcr-9 variant

*Mcr-9* gene was identified by Carroll *et al*. (2019) ^127^ in a colistin susceptible *S.* Typhimurium isolated from a human patient in Washington State in 2010. A BLAST analysis identified the gene in 355 genomes, where 65 were encoded on a plasmid replicon of the same contig ^127^. The presence of *mcr-*9 on a plasmid replicon facilitates its dissemination globally and across *Enterobacteriaceae* ^127^. It has thereafter been identified in more clinical settings ^128–135^ and in food-producing animals ^136, 137^. Among the 355 genomes, 65 were associated with a plasmid replicon, which may facilitate their dissemination globally ^127^. *Mcr-9* cassette is usually located either on a IncHI_2_ and/or IncHI_2A_ replicon or chromosomally encoded ^127, 133, 138, 139^. It is widely distributed within *Enterobacteriaceae*, particularly within *Enterobacter* sp. ^129, 130, 132, 133, 135^, *Citrobacter telavivum* ^134^, *Salmonella* sp. ^127, 137, 138^ and *E. coli* ^136, 140^.

*Mcr-9* is always associated with *wbuC,* encoding a cupin fold metalloprotein, located downstream the gene ^131, 135, 140^. The amino acid sequence of *mcr-9* closely resembles that of *mcr-3* and *mcr-7,* at a 99.5% coverage; it has amino acid sequence identity of 64.5% to *mcr*-3.17 and 63% to *mcr-*7 ^127, 140^. A three-dimensional structural model analysis of all nine *mcr* homologues based on the amino acid sequences of each showed that *mcr-9* was closely related to *mcr-3*, *mcr*-4 and *mcr*-7 at a structural level ^127^. Further, all nine *mcr*-homologues had a high level of conservation for both domains (transmembrane domain and catalytic domain): the structural models showed the amino acids and structural elements were conserved in the C-terminal catalytic domain and the structural elements in the membrane-anchored N-terminal domain ^127^. A database search using *mcr-9* as a template identified a chromosomal-encoded *mcr-9*-like gene within a *Buttiauxella* species isolate, a member of the *Enterobacteriaceae* ^140^. The identified *mcr*-like protein, MCR-BG, was isolated from *B. gaviniae*, and was found to have an 84% amino acid identity to the MCR-9 enzyme; MCR-BG, similar to *mcr-9,* has the *wbuC* gene located downstream ^140^.

Similar results were reported by Yuan *et al.* (2019) with a PEtN transferase isolated from *B. brennerae.* It had 86.83% protein sequence identity to *mcr-9* at a 100% coverage ^135^. Further, the *wbuC* located adjacent the *mcr-9* gene was found to be homologous to that of *Buttiauxella* with a 86% amino acid identity at a 98% coverage ^135^. This evidence suggests that *mcr-9* may have originated from *Buttiauxella* species and disseminated to the rest of the *Enterobacteriaceae* family.

The dissemination of *mcr-9* may have been aided by its association with IS elements flanking the cassette ^133^. The *mcr-9* cassette has been found flanked with IS*903* and IS*903*-like elements upstream as well as IS*1R*, IS*26*-like, and IS*15DI*I located downstream in multiple reports ^132, 133, 135, 140^. The first report of the *mcr-9* cassette encoded the *wbuC* gene, a TCS encoding *qseC* and *qseB* genes, ΔIS*1R* and a remnant of Δ*silR,* encoding a transcriptional regulatory protein, all of which are located downstream of the *mcr-9* gene. The *mcr-9* cassette is thereafter flanked by intact IS*903* elements ^135^. Though two copies of an intact IS element should be able to form a composite transposon or a circular intermediate, an inverse-PCR assay failed to detect any intermediates ^135^. However, the identification of IS*903*-like elements associated with *mcr-9* genes suggests the acquisition of the cassette through an IS*903* dependent mechanism. In some reports, the 3’region of *mcr-9* is adjacent to *copS* (three copper resistance membrane-spanning proteins that includes *rcnA, rcnR* and a domain-containing DUF4942) ^132, 139^.

The *mcr-9* protein as stated above has high levels of conservation within both PEtN transferase domains ^127^. Notwithstanding, multiple reports have shown *mcr-9*-harbouring isolates that are susceptible to colistin ^127, 129, 130^, with the colistin MIC value usually less than 2 mg/L ^128^. Kieffer *et al.* (2019b) ^140^ and Carroll *et al*. (2019) ^127^ performed induction experiments that conferred clinical resistance. However, the mechanism underlining each experiment remains unknown. Carroll *et al.* (2019) identified a conserved σ^70^ family-dependent -35 and -10 regions and an inverted repeat in the promoter of *mcr-9* genes in the database. The authors suggest that the conserved DNA motif in the *mcr-9* promoter is likely a recognition sequence for a transcription regulator, which might be needed for full expression of the *mcr-9* gene ^127^. Kieffer *et al.* (2019b) ^140^ suggested the QseC-QseB TCS played a role in inducing *mcr-9* expression to confer colistin resistance in the presence of sub-inhibitory concentration of colistin; however, multiple reports have failed to replicate these results ^130, 134, 138, 139^. In these studies, the *mcr-9*-harbouring isolates encoding the QseC-QseB TCS regulatory genes were fully susceptible to colistin ^134, 138^. Tyson *et al.* (2020) ^138^ suggests the induction and overexpression of *mcr-9* may be context-dependent and be different in different strain backgrounds. The expression of *mcr-9* in a pBAD vector conferred colistin resistance ^141^ and Kieffer *et al.* (2019b) ^140^ has shown that MCR-9 can mediate Lipid A modifications. However, the genes and molecules required to regulate *mcr-9* expression in its own genomic environment remains unknown ^130^.

### Mcr-10 variant

The *mcr-10* gene was first identified in an *Enterobacter roggenkampii* clinical strain in China ^141^. It has a nucleotide sequence identity and amino acid sequence identity of 79.69% and 82.93% respectively to *mcr-9* ^141^. Therefore, like *mcr-9*, the *mcr-10* gene shares a significant amino acid identity to the chromosomally encoded *mcr*-like PEtN transferase of *Buttiauxella* species ^141^. *mcr-10* encodes a putative PEtN transferase that is made up of an N-terminal membrane-anchored domain and a C-terminal soluble catalytic domain, which have high levels of conservation to both MCR proteins and the MCR-B of various *Buttiauxella* species ^141, 142^. Xu *et al.* (2021) evaluated whether *mcr-10* can mediate colistin resistance and compared its activity to that of *mcr-1* and *mcr-9.* The study showed that the activity of *mcr-10* against colistin is lower than that of *mcr-9* but is able to mediate colistin resistance with an MIC value of 2.5 mg/L ^142^. This shows that *mcr-10* is functional and thus aids the host to survive colistin selective pressure ^142^. Xu *et al.* (2021) again showed that the expression of *mcr-10* is inducible under colistin resistance and further found an upregulation of *phoP-phoQ* TCS genes, producing a colistin MIC value of 8 mg/L. The authors suggested that *mcr-10* might co-function with PhoPQ to mediate high levels of colistin resistance ^142^.

*Mcr-10* has been identified on IncFIA and IncFIB plasmids ^141–144^ and adjacent to a *XerC* gene, which encodes a XerC type tyrosine recombinase found to mobilise adjacent genetic components such as antimicrobial resistance genes ^141^. Thus, *mcr-10* would be presumed to have disseminated across *Enterobacteriaceae* family but has only been identified commonly in *Enterobacter roggenkampii* ^141, 142, 145^ and once in *Cronobacter sakazakii* ^143^. Wang *et al.* (2020a) and Xu *et al.* (2021) suggest that *E. roggenkampii* is an important reservoir of *mcr-10*.

In the initial identification of *mcr-10*, the gene was adjacent XerC and flanked by two intact IS*903* elements which could form a composite transposon with the potential to mediate mobilisation of the *mcr-10* gene ^141^. The analysis of the immediate genetic environment of the IS*903* element found the absence of direct repeats that are generated during an insertion event; thus, the acquisition of the region bracketed by the IS*903* elements was not due to an insertion event ^141^. The presence of an a ΔIS*Ec36* downstream the *mcr-1*0 gene was reported to make it impossible to identify the recombination sites that the XerC-type tyrosine recombinase recognizes ^141^. Thus, the acquisition events of *mcr-10* into p*MCR-1*0_09006 remains unknown ^141^. *Mcr-10* has thereafter been ^145^ identified with various insertion sequences, including complete and truncated remnants of IS*26* and IS*Kpn26*, complete sequences of IS*Ec36* and IS*1*, a ΔIS*Ecl1*-like elements and recently, a new IS element designated IS*Crsa1* ^143, 145^. The identification of various IS elements downstream *mcr-10-xerC* suggests that this region might be a hotspot for insertions of MGEs ^141, 142^. The acquisition of *mcr-10* is presumed to be through site-specific recombination; ^143^ however, evidence of mobilisation of *mcr-10* is scarce.

### Risk factors for acquiring mcr genes

Antimicrobials are commonly administered by farmers to pigs and poultry for prophylaxis, therapeutics, and growth promotion in livestock ^146–148^. Studies around the world have shown that the use of colistin as a growth promoter results in a high frequency of *mcr-1* positive, colistin resistant *Enterobacteriaceae* (MPCRE) because of the high selective pressure in the veterinary environment. The selection pressure led to the acquisition and spread of *mcr-1* genes ^11, 146, 148–152^. The presence of *mcr-1* in food animals had a serious potential to spread MPCRE to humans via foodborne transmission ^11, 150, 151^.

The pharmaceutical form of colistin is indicated for oral administration and is present in a powder or solution form ^152^. Studies have shown that the drug is poorly absorbed in the digestive tract after oral administration and can be excreted in high levels through the faeces ^1, 150, 152^. This results in the presence of colistin or its active metabolites in the environment alongside the MPCRE isolates from the animal faeces ^150^. Xia *et al.* (2019) ^153^ measured the concentration of colistin in animal feeds and fresh manure in five swine farms and found that the average concentration of colistin in animal feed on farm 4 was 60 mg/kg, which was scientifically higher than other farms. The fresh manure samples collected on farm 4 had the highest concentration of 17,383 µg/kg. Other farms had colistin concentrations in their fresh manures ranging from 140 µg/kg in farms 1, 2 and 5, to 7,529 µg/kg in farm 3 ^153^. The researchers also discovered a strong relationship between colistin concentration and the relative abundance of *mcr-1* genes in fresh manure ^153^. The use of this manure as a fertilizer in agriculture or feeding farmed fish can result in the contamination of agriculture and, in some cases, the aquatic environment, polluting rivers and lakes ^150, 154^.

Luo *et al.* (2017) cultured an *mcr-1* producing *Raoultella ornithinolytica*, a member of the *Enterobacteriaceae* family commonly found in soil, aquatic, and botanical environments, in retail vegetables in China. They emphasized that using animal excrements contaminated with colistin and/or MPCRE can cause contamination and spread of MPCRE to fresh vegetables. ^154^. There is, however, no epidemiological data on the contamination of rivers and lakes by MPCRE-contaminated animal excrements and thus, no evidence of the spread of MPCRE to the aquaculture ^150^. However, the presence of *mcr-1*-producing *E. coli* has been reported in duck and fish samples in China. Shen *et al.* (2019) ^155^ found a potential spread of *mcr-1* producing *E. coli* (MPEC) from aquaculture supply chain to humans through aquatic food ^150, 155^. Other examples of the spread of MPCRE into the environment includes the identification and isolation of MPCRE in blowflies (Chrysomya sp.) ^156^ and black kites ^54^. Both cases are of public concern because of the potential transmission into human communities.

Yang *et al.* (2019a) ^156^ showed that blowflies may serve as an environmental reservoir and vector of MPCRE between animals, humans, the environment, and waste (landfills and sewage water) ^156^. The presence of MPCRE in Black kites in Russia highlights the spread of *mcr-1* genes into both wildlife and the environment. Tarabai *et al.* (2019) ^54^ hypothesized that the acquisition of MPCRE was either through contact with the Biysk Municipal landfill or through their food, which was commonly found along the Biya River near their nest. Black kites can thereafter spread MPCRE along their long migratory pathways ^54^. The *mcr-1* gene is commonly found on transferable plasmids, and thus the presence of *mcr-1* producing isolates in the microbiota of insects and wildlife increases the possibility of horizontal transfer of *mcr-1* genes alongside other resistance genes within the microbiota. This increases the environmental gene pool and the spread of multi-drug resistant (MDR) pathogens within insects and wildlife ^156^.

There are therefore multiple reservoirs highlighted above with the potential of spreading towards humans because of the close association between humans and the environment. A study in Vietnam showed that the farmers that are in contact with animals under colistin selection pressure have a high risk of being colonized by MPCRE ^151^. Trung *et al.* (2017) ^151^ found that chickens associated with colistin administration had a high carriage of *mcr-1* producing isolates and the colonization of humans was through exposure to the chickens. The chicken faecal samples had a high prevalence of *mcr-1* (59.4%) and zoonotic transmission prevalence was 34.7% in farmers. Another possible route of transmission of MPCRE from animals under colistin pressure was foodborne transmission ^11, 157, 158^. Monte *et al.* (2017a) found that chicken meat was acting as a reservoir of MPEC. A study performed by Shen *et al.* (2018) ^150^ found a positive correlation between MPEC carriage in the human normal flora and the consumption of farm animals, which included pork and sheep. The correlation analysis also proposed a possible transmission of MPEC from aquatic food to humans ^150^. This data is a single example that highlights the presence of MPCRE in food-producing animals and the transmission route of MPCRE from colistin-exposed animals to humans, which impacts public health care.

This therefore led to the ban on colistin use for growth promotion and disease prevention in the food industry to preserve their effectiveness in human medicine as their use contributes to the rising threat of antibiotic resistance ^159^. Randall *et al.* (2018) ^160^ also demonstrated that discontinuing colistin in farming can result in the elimination of *mcr-1*. This longitudinal study evaluated the presence of *mcr-1* in pig faeces and slurry at two time points ^160^. At the beginning of the study, the majority of the pigs were colonized by MPEC. Twenty months after cessation of colistin use and implementation of hygiene and other measures, MPEC was not detected in samples. These results thus highlight that the use of colistin is the driving force behind the spread of colistin resistance genes and its stability in the population ^160^. The integration of *mcr-1,* however, into conjugative plasmids encoding other resistance genes allows for the use of other antibiotics such as quinolones and extended spectrum cephalosporins to simultaneously co-select for *mcr-1*, allowing for its stability and dissemination ^146, 161, 162^. This, thus, creates severe challenges for controlling the selection and subsequent transmission of *mcr-1* genes ^163^.

After the first detection of *mcr-1* by Liu *et al.* (2016), *mcr-1* genes have been detected in animals, the environment, and humans. Initially, the detection of *mcr-1* genes in humans was associated with the risk factors mentioned above: contact with farm animals, ingestion of contaminated farm food, or fresh vegetables. Epidemiology studies that identified *mcr-1* genes in the clinical settings were accompanied with a questionnaire to identify the epidemiological exposure. In countries such as Finland, where the use of colistin is restricted for rare clinical indications, the first detection of the *mcr-1* gene was identified in a healthy male with previous history of travelling abroad, 6 months before faecal sampling ^164^. Traveling to other countries has been shown to increase the risk of acquiring mcr-1 positive isolates, particularly in patients who consumed meat while traveling abroad ^164, 165^. The use of antibiotics prior to hospitalization or infection has been seen as an important risk factor for a multidrug resistant organism (MDRO) infection ^165^. An epidemiological and clinical study performed by Wang *et al.* (2017) ^166^ discovered that MPEC can acquire other resistance genes, and thus previous use of antibiotics such as carbapenems and fluroquinolones was associated with an increased risk of infection by MPEC ^166^. Thus, the administration of antibiotics in clinics simultaneously promotes the co-selection and the preservation of colistin resistance genes ^167^.

An interesting possible transmission of MPEC is between companion animals and humans. A worker at a pet shop with no prior antibiotic use or travelling abroad was identified carrying an MPEC ^168^. A faecal screening of the pets residing where the man worked identified 4 dogs and 1 cat with an *mcr-1* positive isolate. Zhang *et al.* (2016) ^168^ suggested companion animals as a possible reservoir of colistin-resistant *E. coli*, facilitating the spread of colistin resistance genes within the community.

### Current and future perspectives on treating mcr-mediated colistin resistance

#### Clustered regularly interspaced short palindromic repeats (CRISPR)

Clustered regularly interspaced short palindromic repeats (CRISPR) has been exploited and developed within molecular biology as a site-specific tool for genetic engineering ^169^, and in regards to resistance, as a tool to deliver a programmable DNA nuclease (CRISPR-Cas9) into an MDR-pathogen to eliminate the resistance gene or plasmid ^170^. Dong *et al.* (2019) ^170^ developed a conjugative CRISPR-Cas9 system that aimed to remove plasmids harbouring the *mcr-1* gene from bacteria. The system targeted the *mcr-1* gene and the authors found that the formation of DSBs during the elimination of the resistance gene resulted in the elimination of the whole plasmid in recipient cells ^170^. This thereafter sensitized the recipient cells to colistin. Dong *et al.* (2019) ^170^ further found that the cells are thereafter immune against the *mcr-1* gene. Wang *et al.* (2019a) ^171^ used the same concept but developed the tool to remove both *mcr-1*-harbouring plasmids and MDR plasmids present in recipient cells using partial sequences of the targeted plasmids. He *et al.* (2021) ^172^ used the tool to eliminate both chromosomal and plasmid-borne *mcr-1* genes in *E. coli.* The CRISPR-Cas9 system was designed to target IS*Apl1,* which eliminated *mcr-1* harbouring plasmids, and in the case of chromosomally encoded *mcr-1*, cell death was seen. Similar results were achieved ^170^, where recipient cells further acquired immunity against the acquisition of the exogenous *mcr-1* containing plasmid ^172^. These studies show that CRISPR-Cas9 system is an efficient tool for plasmid curing and to sensitise clinical isolates to antibiotics in vitro ^170, 171^. This tool can be optimized for therapeutic application for the elimination of antibiotic genes from resistance reservoirs such as animal guts with prior exposure to antibiotics, the human microbial flora or the bacteria in natural settings such as wastewater, farms or hospital settings ^170^.

The CRISPR-Cas9 system is however made up of highly anionic nucleic acids and proteins that cannot penetrate the cell membrane into cells and thus requires a delivery vehicle. Sun *et al.* (2017c) ^173^ explored Cathelicidins, which are short cationic antimicrobial peptides, as a plasmid delivery system. The study evaluated the use of BMAP-27 to increase the efficiency of plasmid transfer of the CRISPR-Cas9 system and found that pCas::*mcr* exhibited better, efficient, and specific antimicrobial effects with the help of BMAP-27 ^173^. The CRISPR-Cas9 system is a powerful tool for genome editing, its development into targeting multiple genes with one use ^171^, could aid in the removal of *mcr* variants in resistance reservoirs and in clinical settings to restore colistin activity.

In the meantime, there are multiple studies evaluating existing antibiotics for activity against *mcr*-producing isolates and synergy combinations.

### Novel antibiotics

There are multiple novel antibiotics that have been found to have activity against *mcr-1*-producing isolates, including eravacycline ^6, 174^ and plazomicin ^175^. Eravacycline is a broad-spectrum synthetic tetracycline that has been proven to be effective against clinically important MDR pathogens, including both Gram-negative and gram-positive bacteria ^174^. Fyfe *et al*. (2016) ^6^ found that eravacycline had a bactericidal effect against *mcr-*producing isolates from *Enterobacteriaceae*. The study further showed that although the overexpression of *mcr-1* increased colistin MIC values by 64-folds, there was no effect on eravacycline susceptibility ^6^. Eravacycline activity was thereafter evaluated against 336 isolates collected from hospitals across China and was found to exhibit a good efficacy against all strains ^174^. These isolates included Extended spectrum β-lactamase (ESβL) and carbapenemase-producing *Enterobacteriaceae*, *mcr-*producing *Enterobacteriaceae* and *A. baumannii*, vancomycin-resistant *Enterobacteriaceae*, β-lactamase producing *Haemophilus influenzae,* penicillin-resistant *Streptococcus pneumoniae* and lastly, methicillin-resistant *Staphylococcus aureus* (MRSA), thus showing a positive potential to treat current drug-resistant bacterial infections ^174^. This, however, also puts eravacycline at risk of being overused.

Plazomicin is a novel broad-spectrum semi-synthetic aminoglycoside that was derived from Sisomicin. It has been shown to have activity against a broad spectrum of MDR pathogens, including ESβL- and carbapenemase-producing isolates and fluroquinolone-resistant isolates^175^. Denervaud-Tendon *et al.* (2017) ^175^ evaluated the bactericidal activity of plazomicin against colistin-resistant Enterobacterial strains, which harboured different colistin resistance mechanisms i.e., chromosomal-encoded, *mcr-1*-expressing, and intrinsically resistant strains. Plazomicin activity was further compared with other aminoglycoside antibiotics viz., amikacin, gentamicin, and tobramycin. Plazomicin displayed potent activity against the clinical colistin-resistant Enterobacterial strains irrespective of their resistance mechanisms. However, those that were intrinsically resistant (*Serratia, Proteus, Morganella* and *Hafnia)* had a higher MIC value ^175^.

Denervaud-Tendon *et al.* (2017) also found that amongst all aminoglycoside antibiotics tested, plazomicin was the most potent. They lastly also found that plazomicin’s activity is restricted by aminoglycoside resistance mechanisms such as the 16S rRNA methylase-encoding gene, which resulted in an elevated plazomicin MIC value (>128 mg/L). Plazomicin, however, like other aminoglycosides, has a rapid bactericidal activity with favourable chemical and pharmacokinetics properties and thus could be incorporated into therapeutic treatments against *mcr*-producing bacterial infections ^175^.

Artilysin ® Art-175 is a novel engineered enzyme-based experimental therapeutic derived from endolysins produced by lytic bacteriophages. These endolysins degrade the bacterial cell wall resulting in cell lysis, which occurs at the end of the bacteriophage infection cycle ^176^. Artilysin ® Art-175 is made up of the outer membrane destabilizing peptide, which is fused to an endolysin, it targets the anionic lipopolysaccharide molecules of the outer membrane of Gram-negatives. Schirmeier *et al.* (2018) ^177^ evaluated the potential of Art-175 against pan-drug resistant isolates, including colistin-resistant MPEC and further determined the potential of cross-resistance between colistin and Art-175. Art-175 was highly bactericidal against the tested isolates, which were more susceptible to Art-175 than the colistin-susceptible isolates ^177^. Hence, there is no cross-resistance between the two peptides, highlighting Art-175 as a potent solution against *mcr*-producing isolates ^177^.

### Antimicrobial peptides

Bacteriocins are ribosome-synthesized antimicrobial peptides (AMPs) produced by both Gram-positive and Gram-negative bacteria. Studies have shown that bacteriocins produced by lactic acid bacteria (LAB) are cationic peptides with bactericidal activity that act on the cytoplasmic membrane of susceptible microorganisms ^178^. The LAB bacteriocins create pores in the membrane, resulting in intracellular damage. Al Atya *et al*. (2016) ^4^ evaluated the combinations of nisin and enterocin DD14 LAB bacteriocins with colistin to eradicate colistin-resistant *E. coli* strains in either planktonic states or in biofilm. Whilst colistin and the bacteriocins were ineffective as monotherapy against both planktons and biofilms, colistin-nisin and colistin-enterocin DD14 combination were able to reduce the number of colony-forming units (CFU) of strains in both planktonic and biofilm states. The triple combination of all three molecules, colistin-nisin-enterocin DD14, completely eradicated all *E. coli* strains, including colistin-resistant phenotype ^4^. LAB bacteriocins alone cannot penetrate the outer membrane. However, with the disruption of the LPS via colistin activity, the LAB bacteriocins may thereafter act upon the cytoplasmic membrane ^4^. Al Atya *et al.* (2016) ^4^ therefore highlights LAB bacteriocins as a potential novel adjunctive treatment for colistin-resistant MPEC infections.

There have been multiple reports of AMPs that have been developed for treating MDR pathogens, which have a similar mode of action as colistin ^179^. Van der Weider *et al.* (2019) evaluated the antimicrobial activity of two novel AMPs, AA139 and SET-MM3. AA139 originates from a marine lugworm, *Arenicola marina,* AMP arenicin-3. SET-MM3 is a synthetic tetra-branched peptide linked by a lysine core ^179^. The antimicrobial activity of these AMPs was evaluated against a collection of clinically and genotypically diverse *K. pneumoniae* with different antimicrobial susceptibility profiles. Both AMPs, AA139 and SET-M33, further showed a concentration-dependent bactericidal effect across all isolates, meaning that the susceptibility profile of both AMPs was unaffected by the resistance profile and colistin susceptibility of the isolates ^179^. The AMPs were effective against the colistin-resistant strains, and no cross-resistance was observed between the AMPs and colistin. Thus the antimicrobial activity of AA139 and SET-M33 should further be investigated across other Gram-negatives to evaluate its spectrum of bactericidal activity ^179^.

Cross-resistance between several human cationic AMPs (CAMPs) and polymyxins has been previously reported in PEtN transferase-producing *Haemophilus ducreyi and Campylobacter jeyuni* but rarely in *Enterobacteriaceae*. These CAMPs are part of the intrinsic human immune system and act against Gram-negative bacteria by disrupting the outer and inner membrane through electrostatic interaction, similar to colistin’s mechanism of action^180^. Dobias *et al.* (2017) ^180^ selected a few widely distributed CAMPs in humans, LL-37, α-defensin 5 (HD5) and β-defensin 3 (HDB3) and evaluated whether cross-resistance between defensin 5 (HD5) and β-defensin 3 (HDB3) and evaluated whether cross-resistance between CAMPs and polymyxin would be observed in MPEC. Susceptibility of *E. coli* to the different CAMPs selected varied depending on the CAMP molecule rather than the entire CAMP family. It further showed that the production of MCR proteins did not confer cross-resistance with the human CAMPs selected ^180^.

However, the CAMPs activity studied by Dobias *et al.* (2017), ^180^ compared to other studies ^181^, shows that factors such as bacterial species, level of resistance, and colistin resistance mechanisms may affect the bactericidal levels of CAMPs ^180, 181^. Therefore, although *mcr* production did not affect the bactericidal activity of CAMPs against *E. coli*, it needs to be evaluated across *Enterobacteriaceae* and other Gram-negative bacteria to see how different colistin resistance mechanisms affect the CAMPs’ bactericidal effect. Guachalla *et al.* (2018) ^182^ further evaluated the effectiveness of other derivates of the human immune system against a clinical MPEC strain. This includes humanized monoclonal antibodies that have been previously shown to target the LPS O25b antigen that is associated with *E. coli* ST131-H30 ^182^. The expression of *mcr-1* does not affect the binding of ASn-4 to the LPS’s O-antigen and the integration of the complement membrane-attack complex (MAC) into the LPS-modified outer membrane ^182^. This means the expression of *mcr-1* in *E. coli* ST131-H30 does not affect the functionality of the human immune system, nor does it not aid the strain via immune evasion. The authors thereafter suggested the use of antibodies targeting the LPS’s O-antigen as an alternative strategy to combatting MDR infections against *mcr-1* positive isolates ^182^.

Other novel compounds produced to manage *mcr-*producing isolates are antisense agents with antimicrobial properties; this include phosphorodiamidate morpholino oligomers (PPMOs) and peptide nucleic acids (PNAs) ^183^. These antisense molecules are designed to target mRNA, prevent translation, and restore antimicrobial sensitivity. Daly *et al.* (2017) ^184^ designed peptide-conjugated PPMOs targeted to *mcr-1* mRNA (MCR-1 PPMO) of clinical *mcr-1*-positive *E. coli* strains and showed that these molecules were able to make the strains re-sensitive to polymyxins by *MCR-1* inhibition ^184^. Similar results were achieved by Nezhadi *et al.* (2019) ^183^ with PNAs, designed for inhibition of *mcr-1* translation. They ^183^ showed that the introduction of PNA resulted in a 95% reduction of *mcr-1* expression, measured by RT-PCR, highlighting the efficacy of PNAs ^183^. Daly *et al*. (2017) ^184^ evaluated the antisense approach in a sepsis model using mice and found that a combined therapy of MCR-1 PPMO with colistin reduced morbidity and bacterial burden in the spleen at 24hr. Thus, antisense agents may be effective therapeutics, either alone by targeting an essential gene resulting in cell death, ^184^ or in combination with colistin ^183, 184^ to treat *mcr-1*-producing isolates. However, these molecules have varying toxicity effects and more *in-vivo* research is required prior to clinical application ^183^.

### Natural compounds

Other novel compounds that have been reported to restore susceptibility of polymyxins in *mcr-1* producing isolates includes the novel *MCR-1* inhibitor, Osthole (7-methoxy-8-(3-methyl-2-buteryl) coumarin. Osthole (OST) is a natural compound derived from the dried root and rhizome of *Cnidium monnieri* ^185^. Zhou *et al.* (2019) ^185^ evaluated the effects of OST on the inhibition of *MCR-1* enzyme and the mechanisms behind the inhibition. The colistin-OST combination had a synergistic effect as a mouse-thigh-infection model showed that the combination exhibits a bactericidal activity against *mcr-1*-producing *Enterobacteriaceae,* significantly reducing the bacterial load in the thighs following subcutaneous administration ^185^. Using a molecular dynamic stimulation, Zhao *et al.* (2019) ^185^ found that OST could localize in the binding pocket (residue 330-350) of MCR-1, blocking the substrate and reducing MCR-1’s biological activity. OST therefore inhibits MCR-1 in *Enterobacteriaceae,* making colistin potent ^185^. Due to the conservation of MCR proteins, OST may have the same inhibition efficiency across the other MCR variants.

Natural/organic compounds identified with a synergistic effect on colistin include Honokiol ^186^, Isoalantolactone (IAL) ^187^, Calycosin ^188^ and Eugenol ^189^. Honokiol was derived from the dry skin and bark of *Magnolia officinalis* ^186^, IAL is the main sesquiterpene lactone from Radix inulae and other plants ^187^, Calycosin is a flavonoid from a direct root extract of the traditional Chinese medicinal herb, Radix astragali.^188^ Lastly, Eugenol is a phenylpropanoid, an essential oil isolated from plants ^190^. Each of these compounds, when combined with colistin, decreased colistin MICs and increased colistin’s bactericidal activity against colistin-resistant isolates. Eugenol decreased the expression of *mcr-1* ^189^ and honokiol bound directly to the active site of MCR-1, inhibiting its activity ^186^. Guo *et al*. (2020) ^186^ and Liu *et al*. (2020) ^188^ showed that the natural compounds, honokiol and calycosin, in combinations with colistin, respectively reduced the load of bacteria and improved viability in animal models.

### Efflux pump inhibitors

Efflux pumps play a role in reducing colistin susceptibility in *Enterobacteriaceae* ^10^. Baron *et al*. (2018) ^191^ found that an efflux pump inhibitor, carbonyl cyanide 3-chlorophenylhydrazone (CCCP), was a good alternative to reverse colistin resistance in colistin-resistant isolates irrespective of their molecular resistance mechanisms. The study shows that CCCP was able to restore colistin activity across a diverse set of strains carrying different colistin resistance mechanisms (*mcr-1*, *pmrAB*, *mgrB*, etc) ^191, 192^. In *mcr-1* producing isolates, it was seen that colistin-CCCP combination inhibited the transcription of the *mcr-1* gene. The mechanism behind this observation, however, is unknown ^191^.

### FDA-approved drugs

The food and drug administration (FDA) has a library of FDA-approved drugs that can be screened for bactericidal activity on *mcr*-producing *Enterobacteriaceae* isolates (Prestwick Chemical, Illkirch-Graffenstuden, France). Multiple compounds such as pentamidine, zidovudine ^193^, sulphonamide compounds ^193^, a polymyxin derivative, NAB739, ^194^ and compound PFK-185, ^194^ have been identified during a library search.

Zidovudine is a nucleoside reverse transcriptase inhibitor. In the 1980s, it was used as an anticancer drug ^195^. In 1986, its antibacterial effect was revealed ^196^ and in 1987, it was used as the first antiretroviral for treatment against HIV infections ^195, 197^. Peyclit *et al*. (2018) ^197^ investigated the antibacterial activity of Zidovudine against a large number of characterised MDR *Enterobacteriaceae* strains isolated from different geographical areas, including carbapenem- and colistin-resistant isolates; Zidovudine was effective against *Enterobacteriaceae*. Despite their antimicrobial susceptibility profile, the MIC values ranged between 0.05-1.67 µg/mL ^197^.

Another antiretroviral drug that is used to treat HIV/AIDS but also has bactericidal activity against Gram-negative bacteria is Azidothymidine (AZT). Hu *et al.* (2019) ^198^ shows that AZT, in combination with colistin, was able to eradicate MDR *Enterobacteriaceae* strains with different antimicrobial susceptibility profiles and enhance the activity of colistin. The combined therapy was also effective therapeutically in a murine peritoneal infection against NDM-1-producing *K. pneumoniae* and MPEC ^198^.

Further, Pentamidine is an antiprotozoal drug that Stokes *et al*. (2017) ^199^ identified as an effective antibiotic adjuvant producing synergistic combination activity against a wide range of Gram-negative bacteria. The adjuvant was identified during a library screening of non-lethal, outer membrane-active compounds ^199^. The study shows that Pentamidine directly associates with the outer membrane by inhibiting the oligosaccharide (OS) core biosynthesis, releasing LPS from the outer membrane ^199^. Stokes *et al.* (2017) ^199^ further showed that Pentamidine has synergistic activity with hydrophobic antibiotics such as rifampicin, novobiocin, and erythromycin but not with hydrophilic and low-molecular weight antibiotics ^199^. Although the study does not evaluate synergistic combinations with Pentamidine, Stokes *et al.* (2017) ^199^ concluded that pentamidine can be used as an antibiotic adjuvant for treating colistin-resistant infections.

Barker *et al.* (2017) ^200^ screened a diverse cohort of adjuvants to identify a compound that could both sensitise colistin-resistant and hypersensitise colistin-susceptible bacteria to colistin, to potentially lower the effective dosage of colistin, reducing its toxicity. The study identified three compounds that were potent modulators of colistin resistance and were able to significantly reduce MIC values in both plasmid-and chromosomal-mediated resistance mechanisms, and further hypersensitize colistin-susceptible isolates ^200^

Another compound identified in the FDA-approved library with synergistic activity with colistin is sulfadiazine (SDI), a sulphonamide compound. Okdah *et al.* (2018) ^193^ evaluated the potential of different sulphonamide compounds with potential synergistic and bactericidal activity in combination with colistin; SDI had the highest synergistic effect. The combination of SDI-colistin was effective, independent of colistin resistance mechanism, across the broad range of MDR bacterium strains tested ^193^.

Zhang *et al.* (2019) ^194^ identified an antitumor drug, PFK-185 and its analogs, PFK-015 and 3PO, during a screening of the clinical compound library. These compounds can exert synergise with colistin against colistin-resistant *Enterobacteriaceae* despite *mcr* expression and the antimicrobial susceptibility profile ^194^. Zhang *et al.* (2019) ^194^ found that PFK-185 had no effect on cellular morphology when used alone. However, when in combination with colistin, it enhanced the bacterium-killing effect of colistin, increasing the survival rate to 60%. The combined colistin-PFK-158 therapy had the most significant bactericidal activity, with the most significant reductions in the bacterial burdens post-*in vivo* experiments and during time-kills studies. ^194^.

The last compound reported from the FDA-approved library is a novel polymyxin derivative, NAB739, which carries only three of the five amino groups of colistin, each placed in a strategic position ^201^. Tyrrell *et al.* (2019) ^201^ showed that NAB739 sensitizes polymyxin-resistant strains to rifampicin and a combination of the two was synergistic against ten of the eleven colistin-resistant strains. Tyrrell *et al.* (2019) ^201^ further showed that NAB739 was also synergistic with meropenem and retapamulin. As well, NAB739 in combination with other compounds has significant bactericidal activity against polymyxin-resistant isolates and compared to polymyxins, it has a better tolerability and efficacy ^201^. Thus, NAB739 combination therapies may replace polymyxin for treating MDR pathogens ^201^.

These studies ^197, 199^ however, show that old antimicrobials listed within the FDA-approved library may be re-introduced against bacteria that are resistant to multiple antibacterials in current use ^197^.

### Combination therapy

There are multiple compounds, either antibiotics or adjuvants, which are synergistic with colistin, thus reducing the dosage of colistin required for treatment, minimizing its toxicity, overcoming colistin resistance, and retaining maximal therapeutic efficacy ^198, 200^. Colistin activity has been restored when it is combined with antibiotics such as rifampicin ^202, 203^, rifabutin, minocycline ^203^, amikacin ^204^, tigecycline ^205^ and clarithromycin ^206^. In all these reports, the antibiotics were ineffective as monotherapy against MPCRE isolates. However, in combination with colistin, they resulted in significant bactericidal activity. Yu *et al.* (2019) ^203^ found that colistin in combination with rifampicin, rifabutin or minocycline was able to eradicate XDR, NDM- and *mcr–* co-producing *E. coli in-vitro* and in mouse models. Other combinations that were confirmed effective against *mcr-1*-producing colistin-resistant *E. coli* using an animal model include tigecycline-colistin ^205^, clarithromycin-colistin ^206^ and a triple combination of colistin-rifampin-azithromycin discovered by Li *et al.* (2018b) ^202^. Another triple combination therapy that was discovered was colistin-aztreonam-amikacin, which was effective against both carbapenemase-producing and *mcr -1*-producing *Enterobacteriaceae* isolates ^207^.

Lastly, a combination therapy that was effective against both carbapenem- and colistin-resistant *Enterobacteriaceae* isolates was the β-lactam-β-lactamase inhibitor, imipenem-relebactam, combination ^208^. This combined therapy was potent against colistin-resistant carbapenemase-producing isolates, making it a potential agent against carbapenemase-producing *Enterobacteriaceae;* particularly, those that are colistin resistant (irrespective of the mechanism of resistance) ^208^.

## Conclusion

Herein, we show that *mcr* genes are commonly identified in *E. coli*, *K. pneumoniae,* and *Salmonella* sp., with clones within these strains being disseminated globally within animals, food, the environment, and humans. *Mcr* genes were initially identified in animal samples, specifically livestock animals that were treated with colistin as a therapeutic against Gram-negative bacterial infections or as a growth promoter. *Mcr* has spread to the environment through contaminated but untreated animal faeces used as manure and through contaminated food-producing-animals’ products on markets. Contact with these colistin-treated livestock or their faeces by humans was an identified transmission route. Human specimens currently have the highest reports of *mcr* genes due to the multiple routes of transmission from livestock, the environment and from food, and the use of colistin as a therapeutic against carbapenem-resistant *Enterobacteriaceae* infections. The presence of clones such as *E. coli* ST744 or ST101 in all sources, including animals, the environment, food, and humans, highlights these transmission routes. This makes *mcr* a concerning antibiotic resistance gene of great interest and priority.

Further, *mcr* genes are commonly associated with mobile genetic elements. IS*Apl1* elements have facilitated the horizontal transfer of *mcr*-1 within cassettes of composite transposons. They have been also associated with IncX_4_ plasmids, facilitating their horizontal and vertical transfer across species, genera and families.

Although the *mcr variants* have different possible progenitors viz., *Moracella* sp., *Aeromonas* sp., and *Shewanella* sp. for *mcr-1*, *mcr-3* and *mcr-4,* respectively, the MCR proteins are very well conserved. Each of the 10 MCR proteins encodes a two-domain integral membrane protein with a C-terminal periplasmic domain and an N-terminal 5’-helix transmembrane domain, each MCR protein further encodes the five conserved residues i.e., E248, T286, H389, D458, and H359, located within the active site. PEtN transferase activity was found for each MCR protein, and each was able to mediate colistin resistance, although *mcr-9* expression requires a specific genetic environment to regulate its expression. The conservation seen within MCR proteins, allows for the use of similar therapies for the management of MCRPE isolates, specifically in the cases of identified MCR-1 inhibitors. Research on novel therapeutics is well-summarised in this review, with techniques such as CRISPR-Cas9, peptide nucleic acids, and antimicrobial peptides, which eliminate *mcr* genes within the host. Other novel therapeutics were identified by reviving old FDA-approved drugs. Some were effective against *mcr-*producing isolates or were synergistic with colistin.

These therapeutics, however, need to be further evaluated for their toxicity in humans. This will aid in alleviating the threat imposed by MCRPE in the public health sector

## Data Availability

All data are attached as supplementary files

## Funding

This work was funded by a grant from the National Health Laboratory Service (NHLS) given to Dr. John Osei Sekyere under grant number GRANT004 94809 (reference number PR2010486).

## Acknowledgements

None

## Transparency declaration

**None**

## Notes

### Competing Interest Statement

The authors have declared no competing interest.

### Funding Statement

This work was funded by the NHLS development grant given to John Osei Sekyere

### Author Declarations

All data used for this work are publicly available and also attached as supplementary files

